# Trends in Nationally Notifiable Infectious Diseases in Humans and Animals During the COVID-19 Pandemic in South Korea

**DOI:** 10.1101/2023.10.16.23297064

**Authors:** Taehee Chang, Sung-il Cho, Dae sung Yoo, Kyung-Duk Min

**Affiliations:** Department of Public Health Sciences, Graduate School of Public Health, Seoul National University, Seoul, Republic of Korea; Institute of Health and Environment, Seoul National University, Seoul, Republic of Korea; College of Veterinary Medicine, Chonnam National University, Gwangju, Republic of Korea; College of Veterinary Medicine, Chungbuk National University, Republic of Korea

**Keywords:** Non-pharmaceutical intervention, COVID-19 pandemic, Infectious disease, ARIMA model

## Abstract

Non-pharmaceutical interventions (NPIs) were implemented to cope with the coronavirus disease 2019 (COVID-19) pandemic in South Korea. These interventions could also have affected other infectious diseases, but there have been no comprehensive studies regarding their impacts. This study examined trends in notifiable infectious diseases in both humans and animals during the COVID-19 pandemic. Autoregressive integrated moving average (ARIMA) models were developed for each disease using data from 2016 to 2019, and the incidences for 2020 to 2021 were predicted. Subsequently, the predicted numbers of cases were compared with actual observations. Our findings indicated a substantial reduction in human respiratory infectious diseases during implementation of NPIs. However, human gastrointestinal infectious diseases and livestock diseases did not show a significant decrease. The results revealed that the preventive effect sizes of NPIs varied among diseases and indicated the potential for side effects, suggesting that complementary interventions are needed to minimize these negative effects.

## 1. Introduction

The global coronavirus disease 2019 (COVID-19) pandemic, caused by severe acute respiratory syndrome coronavirus 2 (SARS-CoV-2), has dramatically disrupted the lives of people around the world, resulting in record numbers of both cases and fatalities [1]. In the early stages of the pandemic, the major public health measures were non-pharmaceutical interventions (NPIs) such as social distancing, mask-wearing, and contact tracing. NPIs have proven effective in mitigating the epidemic curves in various contexts, even without vaccines or specific treatments targeting the pathogen [2–4]. Since March 2020, stringent public health measures have been implemented nationwide in South Korea, effectively suppressing the spread of COVID-19 [5–7].

The effects of NPIs are not necessarily limited to COVID-19, and they could influence the incidences of other infectious diseases. Because the major mechanism underlying the effects of NPIs comprises reducing effective contact within the population, NPIs can also mitigate other respiratory infectious diseases [5, 8, 9]. Furthermore, with the implementation of social distancing measures (e.g., restrictions on social gatherings in restaurants) and improved personal hygiene practices, it is possible to reduce the occurrence of gastrointestinal diseases [9, 10]. The effects of COVID-19 can also extend beyond human diseases and affect the risks of infectious diseases in animals [11–13]. Human movement restrictions and the global economic crisis have significantly disrupted farming operations, veterinary services, wildlife surveillance, and zoonotic disease control, with broad impacts on animal health and welfare [11, 12]. These adverse effects can contribute to the outbreak of major zoonotic diseases, such as brucellosis and bovine tuberculosis in animal populations, increasing the risk of zoonotic spillover [13].

Several studies have investigated the effects of NPIs implemented during the COVID-19 pandemic on other infectious diseases in South Korea. Some have shown that reductions in respiratory infection trends coincided social distancing interventions [14–20]. However, the effects of NPIs on gastrointestinal diseases are inconsistent. For example, a notable reduction in viral gastrointestinal infections was reported, but there were no significant decreases in bacterial infections such as *Campylobacter* spp., *Clostridium perfringens*, and *Salmonella* spp. [10, 18, 20]. This difference presumably occurred because viral gastrointestinal diseases are primarily transmitted through fecal–oral contamination or direct contact between individuals. In contrast, bacterial gastrointestinal infections mainly occur as foodborne illnesses through the consumption of contaminated food or water [21]. Therefore, there is a need for research that can consolidate fragmented phenomena, focusing on nationally notifiable infectious diseases in humans and livestock, using data collected post-2020.

This study was performed to examine trends in the incidences of notifiable infectious diseases in humans and animals during the period of COVID-19-related social distancing in South Korea. We built time series models [22] for six respiratory human infectious diseases (varicella, pertussis, mumps, invasive pneumococcal disease, scarlet fever, and tuberculosis), four human gastrointestinal diseases (typhoid fever, shigellosis, hepatitis A, and enterohemorrhagic *Escherichia coli*), and two livestock diseases (cattle tuberculosis and cattle brucellosis). Considering the decreased stringency of NPIs beginning in the first half of 2022 and the potential attenuation of their effects, we focused on the period from 2016 to 2021 in this study.

## 2. Methods

### 2.1 Study design

This study retrospectively analyzed the impact of implementing NPIs for COVID-19 on the incidences of infectious diseases in South Korea. The criteria used to select target infectious diseases from among nationally notifiable diseases were as follows: the principal mode of transmission is respiratory (airborne or droplet) or gastrointestinal (foodborne or fecal–oral route); animal infectious diseases with a risk of zoonotic transmission were selected; and the annual average incidence was > 100 cases. The pre-intervention period was defined as January 2016 to February 2020, and the intervention period was defined as March 2020 to December 2021. From May 2022 onward, the outdoor mask mandate was conditionally lifted. Thus, data up to 2021 were utilized to accurately assess the impact of NPIs.

We developed models utilizing an autoregressive integrated moving average (ARIMA) model to forecast incidences during the intervention period based on patterns in the pre-intervention period. The predicted values were compared with the observed values from 2020 to 2021. Then, visual analysis was performed to assess whether the observed incidences were within the 95% prediction intervals of the predicted values. The reproduction number provides a better understanding of the transmission dynamics of respiratory infectious diseases [23]. Therefore, we calculated and utilized the reproduction number for respiratory infectious diseases in the time series forecasting process. Although tuberculosis is a respiratory infectious disease, we did not calculate its reproduction number because of its complex transmission routes and long latent period; instead, we utilized reported cases for time series forecasting of tuberculosis.

The results of previous studies regarding the impacts of COVID-19 and NPIs on other diseases suggested that the reduced burdens of target diseases during the early stages of the COVID-19 pandemic could be attributed to pandemic-related decreases in health care utilization and disease diagnoses [19]. Therefore, to adjust for the impact of health care utilization, we collected information about annual hospital visits [24] and annual health insurance claims (Table S1) [25], then used those numbers as denominators for disease incidence. In addition, to calculate the incidence rate per population, we collected annual midyear population data for each year in Korea [26]. Total annual medical expenses for each infectious disease were collected to evaluate the impacts of changes in disease occurrence after NPI implementation on the overall disease burden [27]. We calculated each year’s medical expenses per case using Health Insurance Review and Assessment Service (HIRA) data from 2018 to 2021. Then, we multiplied these expenses per case by the estimated and observed cases for each disease to determine the model-based medical costs and observation-based values for each disease. We compared these values to assess changes in overall disease burden.

### 2.2 Social distancing measures

In February 2020, in response to the COVID-19 outbreak in China, South Korea implemented a universal mask mandate and recommended physical distancing. After the increase in COVID-19 cases in South Korea, nationwide social distancing requirements were implemented with various restrictions starting in March 2020 [28]. During the initial phase of the COVID-19 pandemic, the “Distancing in Daily Life” strategy was implemented [29]. After multiple outbreaks occurred near metropolitan areas, the “Distancing in Daily Life” strategy was restructured on June 28, 2020, into a three-tier social distancing system that consisted of Levels 1, 2, and 3 (Table S2) [1]. In November 2020, the social distancing system was reorganized into a five-tier structure that consisted of Levels 1, 1.5, 2, 2.5, and 3 (Table S3). Subsequently, in July 2021, the system was modified to a four-tier structure that consisted of Levels 1, 2, 3, and 4 (Table S4) [29]. In this study, the policy changes were documented based on the most current four-tier structure; any rapid changes within short periods (e.g., 1–2 weeks or 1 month) were not considered because they may not have shown sufficient effectiveness (Fig. 1).

**Fig. 1.**
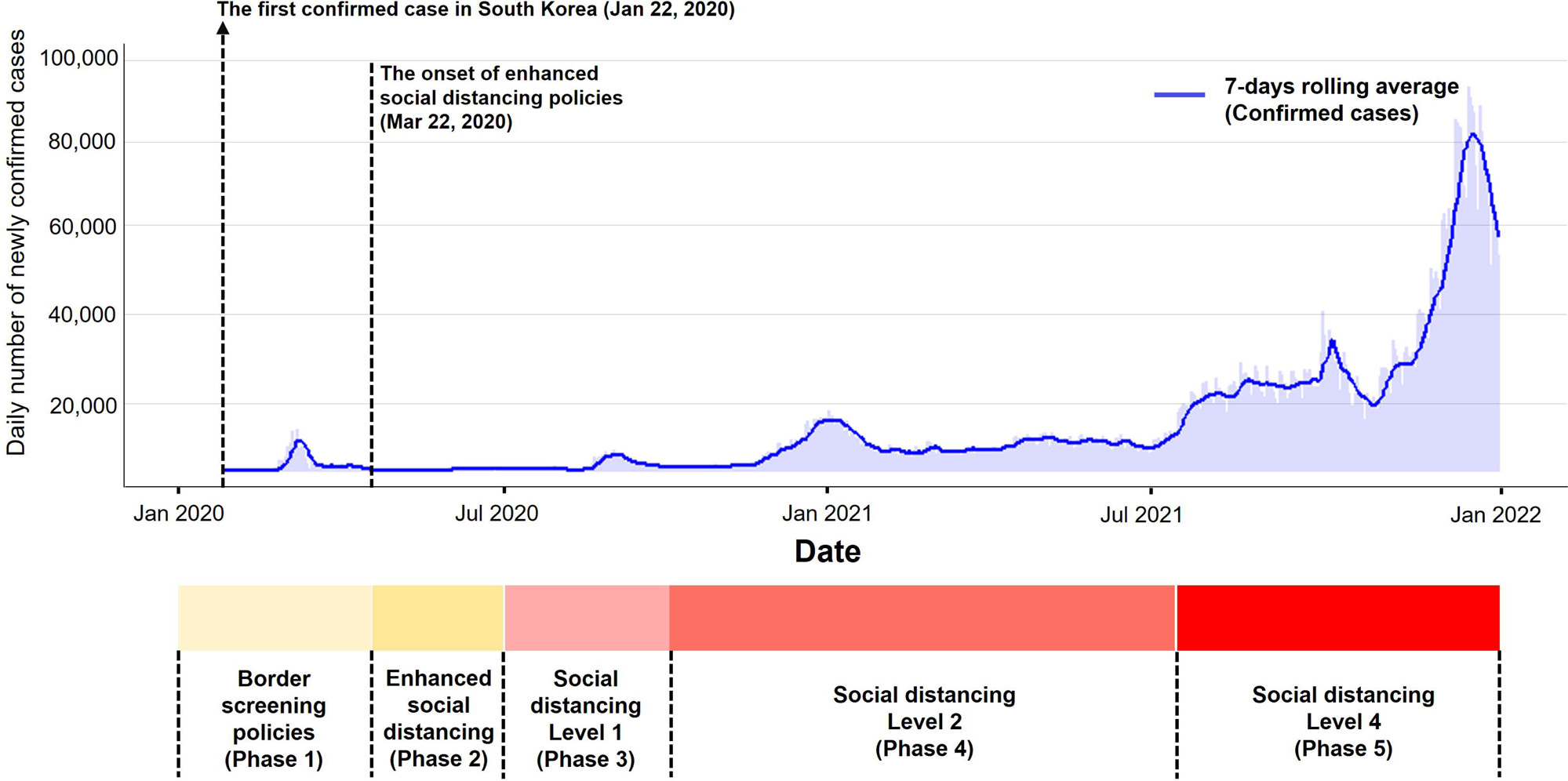
Daily number of confirmed cases and 7-day rolling average number of COVID-19 cases. Levels of NPIs are based on the four-tier system implemented in July 2021.

### 2.3 Data acquisition

We collected data regarding weekly and monthly domestic cases of nationally notifiable infectious diseases from the Infectious Disease Portal [30] provided by the Korea Disease Control and Prevention Agency (KDCA). To minimize sampling bias and perform more robust analysis considering the COVID-19-related decrease in health care utilization, we solely focused on infectious diseases under the mandatory surveillance system. Therefore, we collected records of six respiratory infectious diseases (varicella, pertussis, mumps, invasive pneumococcal disease, scarlet fever, and tuberculosis) and four gastrointestinal diseases (typhoid fever, shigellosis, hepatitis A, and enterohemorrhagic *E. coli*) from February 2016 to December 2021 (Table S5).

Data were collected from the Korea Animal Health Integrated System (KAHIS) to investigate animal diseases with zoonotic potential [31]. KAHIS is a comprehensive system operated by the Animal and Plant Quarantine Agency (APQA) that integrates and provides nationwide information about livestock diseases. Therefore, we selected two livestock diseases (cattle tuberculosis and cattle brucellosis) with potential zoonotic infection routes and collected disease occurrence information from January 2016 to December 2021 (Table S5). In addition, we collected data regarding the annual number of livestock and annual scale of livestock farming [32] to calculate the incidence rate relative to the livestock population.

### 2.4 Reproduction number estimation

The reproduction number, used for analysis of respiratory infectious diseases except tuberculosis, is expressed as shown below [23]:

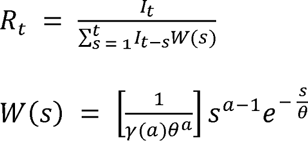

where *R_t_* is the reproduction number, *t* is the number of days elapsed since the start of the epidemic, *I*_t_ is the number of cases on day *t*, *W(s)* is the current infectivity on day *s* after infection, *a* is the shape parameter, and θ is the scale parameter. To estimate current infectivity *W(s)*, we utilized the serial interval and standard deviation for each disease (Table S6) [33–35].

### 2.5 Time series analysis

The ARIMA (p, d, q) model is a time series forecasting technique that incorporates elements of autoregressive (AR), moving average (MA), and AR+MA models to make predictions [22]. ARIMA is commonly used to predict short-term impacts and trends of acute infectious diseases [9, 22]. The parameters p, d, and q in the ARIMA model indicate the order of autoregression, the degree of differencing applied to the original time series, and the order of moving averages, respectively. With respect to some infectious diseases that exhibit seasonality (especially respiratory diseases), we used a seasonal ARIMA (SARIMA [p, d, q] [P, D, Q] s) model. In SARIMA, the additional parameters P, D, Q, and s correspond to seasonal autoregression, seasonal integration, seasonal moving average, and seasonal period length, respectively. Time series forecasting based on the Box-Jenkins method consists of four steps: identification, estimation, diagnostic checking, and forecasting [36]. We adhered to these steps when making predictions.

All data processing and analyses were performed using R software (v. 4. 2. 2) [37]. The *forecast* package [38] was used to model species niches. The corresponding R code is publicly available (https://github.com/TaeHChang/For-the-Paper-4).

## 3. Results

### 3.1 Incidences of human respiratory diseases

This study included six respiratory infectious diseases (Table S5). After the implementation of nationwide social distancing measures in March 2020 (Fig. 1), there were considerable and statistically significant decreases in the weekly reported case numbers (Table 1). The mean weekly incidences for 2016–2019 per 1 million population varied for each disease as follows: varicella, 30.11; pertussis, 0.18; mumps, 6.95; invasive pneumococcal disease, 0.21; scarlet fever, 5.57; tuberculosis, 13.13. However, after implementation of NPIs, the mean weekly incidences for 2020–2021 substantially decreased, with slight variations among phases: varicella, 12.09; pertussis, 0.05; mumps, 4.00; invasive pneumococcal disease, 0.13; scarlet fever, 0.80; tuberculosis, 9.16. The annual medical expenses related to respiratory infectious diseases decreased by 3.77% in 2020, compared with the value calculated using the average estimated incidence; the value decreased by an additional 18.91% in 2021 (Table 2). Whereas medical expenses related to respiratory infectious diseases showed an overall decreasing trend, tuberculosis-related expenses showed a slight increase in 2020; scarlet fever-related expenses also exhibited a slight increase in 2021.

**Table 1.** Weekly Average Incidences of Notifiable Infectious Diseases Included in the Study.

| **Human Diseases**<br>**(per million population)** | 2016-2019 | 2020-2021 | Phase 1<sup>*</sup> | Phase 2<sup>†</sup> | Phase 3<sup>‡</sup> | Phase<br>4<sup>§</sup> | Phase 5<sup>¶</sup> |
| --- | --- | --- | --- | --- | --- | --- | --- |
| **Respiratory diseases** |  |  |  |  |  |  |  |
| Varicella | 30.11 | 12.09 | 25.76 | 9.29 | 9.31 | 8.27 | 7.81 |
| Pertussis | 0.18 | 0.05 | 0.17 | 0.03 | 0.01 | 0.01 | 0.01 |
| Mumps | 6.95 | 4.00 | 3.89 | 4.28 | 4.34 | 3.29 | 4.21 |
| Invasive<br>pneumococcal disease | 0.21 | 0.13 | 0.28 | 0.11 | 0.08 | 0.1 | 0.1 |
| Scarlet fever | 5.57 | 0.80 | 2.12 | 0.76 | 0.59 | 0.34 | 0.21 |
| Tuberculosis | 13.13 | 9.16 | 10.02 | 9.17 | 9.31 | 8.94 | 8.34 |
| **Gastrointestinal or**<br>**enteroviral diseases** |  |  |  |  |  |  |  |
| Typhoid | 0.04 | 0.03 | 0.03 | 0.05 | 0.04 | 0.02 | 0.03 |
| Shigellosis | 0.02 | 0.02 | 0.03 | 0.04 | 0.01 | 0.01 | 0.01 |
| Hepatitis A | 3.02 | 1.91 | 1.37 | 1.58 | 1.59 | 2.48 | 2.53 |
| Enterohemorrhagic *E. coli*<br>infections | 0.05 | 0.12 | 0.04 | 0.23 | 0.21 | 0.06 | 0.06 |
| **Veterinary diseases**<br>**(per 100,000 numbers)** |  |  |  |  |  |  |  |
| Bovine Tuberculosis | 2.19 | 1.29 | 1.04 | 1.82 | 1.43 | 1.27 | 0.91 |
| Bovine Brucellosis | 0.49 | 0.61 | 0.41 | 0.51 | 0.55 | 0.83 | 0.74 |

199 <sup>\*</sup>20 Feb 2020 ~ 21 Mar 2020200 <sup>†</sup>22 Mar 2020 ~ 27 Jun 2020201 <sup>‡</sup>28 Jun 2020 ~ 22 Aug 2020202 <sup>§</sup>23 Aug 2020 ~ 26 Jul 2021203 <sup>¶</sup>27 Jul 2021 ~ 31 Dec 2021

**Table 2.** Annual medical expenses due to infectious diseases (the unit is one million USD).

|  | 2018 | 2019 | 2020<br>lower 95 % | 2020<br>average | 2020<br>upper 95 % | 2020<br>observed (%)<sup>*</sup> | 2021<br>lower 95 % | 2021<br>average | 2021<br>upper 95 % | 2021<br>observed (%)<sup>†</sup> |
| --- | --- | --- | --- | --- | --- | --- | --- | --- | --- | --- |
| **Respiratory**<br>**diseases** |  |  |  |  |  |  |  |  |  |  |

|  |  |  |  |  |  |  |  |  |  |  |
| --- | --- | --- | --- | --- | --- | --- | --- | --- | --- | --- |
| Varicella | 5,399 | 5,590 | 3,704 | 7,153 | 13,548 | 2,675 (-62.61) | 2,692 | 6,559 | 16,001 | 1,971 (-69.95) |
| Pertussis | 544 | 298 | 44 | 380 | 1,171 | 42 (-88.98) | 13 | 409 | 1,900 | 6 (-98.65) |
| Mumps | 1,283 | 1,445 | 1,203 | 1,552 | 2,189 | 790 (-49.08) | 1,066 | 1,532 | 2,203 | 775 (-49.42) |
| Invasive pneumococcal disease | 1,988 | 1,686 | 1,305 | 2,920 | 6,437 | 1,360 (-53.41) | 1,305 | 3,370 | 8,717 | 1,116 (-66.87) |
| Scarlet fever | 1,283 | 691 | 182 | 465 | 947 | 317 (-31.80) | 28 | 124 | 567 | 154 (24.27) |
| Tuberculosis | 61,241 | 63,535 | 42,695 | 48,705 | 55,593 | 53,681 (10.21) | 48,536 | 59,956 | 74,117 | 54,324 (-9.39) |
| **Subtotal** | 71,738 | 73,244 | 49,134 | 61,174 | 79,886 | 58,864 (-3.77) | 53,640 | 71,951 | 103,505 | 58,345 (-18.91) |
| **Gastrointestinal diseases** |  |  |  |  |  |  |  |  |  |  |
| Typhoid | 153 | 41 | 6 | 17 | 62 | 27 (89.81) | 5 | 22 | 102 | 41 (90.625) |
| Shigellosis | 36 | 49 | 6 | 22 | 40 | 12 (-46.86) | 3 | 21 | 126 | 15 (-28.06) |
| Hepatitis A | 2,248 | 27,297 | 1,477 | 6,193 | 22,857 | 4,820 (-22.17) | 1,466 | 9,407 | 60,328 | 9,769 (3.85) |
| Enterohemorrhagic *E.coli* infections | 120 | 157 | 94 | 176 | 275 | 364 (106.61) | 86 | 184 | 388 | 244 (33.06) |
| **Subtotal** | 2,557 | 27,544 | 1,583 | 6,408 | 23,234 | 5,223 (-18.49) | 1,560 | 9,633 | 60,945 | 10,070 (4.53) |
| **Total** | 74,295 | 100,788 | 50,717 | 67,582 | 103,120 | 64,087 (-5.17) | 55,201 | 81,584 | 164,450 | 68,415 (-16.14) |

\*The change from the average estimates of the year 2020
†The change from the average estimates of the year 2021

ARIMA models were constructed; the parameters, diagnostic plots, and model characteristics are presented in the Supplementary materials (Table S7–13, Fig. S1–12). Except for tuberculosis, the actual incidences of diseases examined during the intervention period were significantly lower than the predicted incidences (Fig. 2A–J). The incidences of tuberculosis were lower than predicted values but fell within the 95% prediction intervals of the predicted values. Though they showed a significant decrease compared with the predicted values after implementation of social distancing measures, but no significant difference was observed from the second half of 2020 (Fig. 2K, L).

**Fig. 2.**
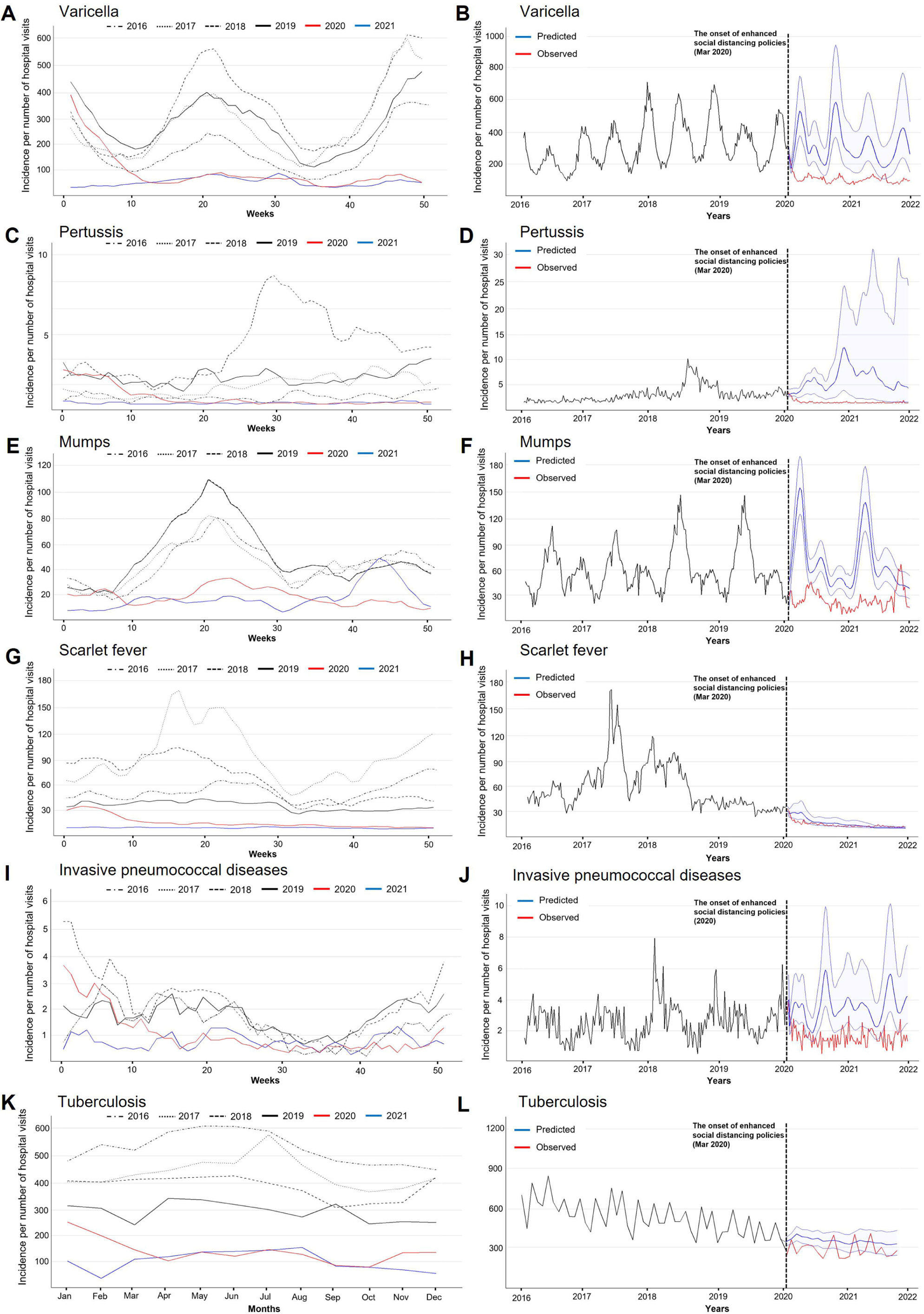
A, C, E, G, I, Weekly incidences of respiratory infectious diseases from the national surveillance system for notifiable infectious diseases, South Korea, 2016–2019 vs. 2020– 2021. B, D, F, H, J, Observed and predicted weekly incidences from 2016 to 2021. K, Monthly incidences of tuberculosis. L, Observed and predicted weekly incidences of tuberculosis.

### 3.2 Incidences of human gastrointestinal diseases

This study included four gastrointestinal infectious diseases (Table S5). Unlike respiratory infectious diseases, the incidences of gastrointestinal diseases did not show significant decreases after implementation of NPIs (Table 1). The mean weekly incidences for 2016– 2019 per 1 million population varied among diseases: typhoid, 0.04; shigellosis, 0.02; hepatitis A, 3.02; enterohemorrhagic *E. coli* infection, 0.05. Although there were slight variations among phases, the mean weekly incidences for 2020–2021 after the implementation of social distancing measures were as follows: typhoid, 0.03; shigellosis, 0.02; hepatitis A, 1.91; enterohemorrhagic *E. coli* infection, 0.12. The annual medical expenses related to gastrointestinal infectious diseases decreased by 18.49% in 2020, compared with the value calculated using the average estimated incidence; the value increased by 4.53% in 2021 (Table 2). The trend in medical expenses related to gastrointestinal infectious diseases varied according to specific conditions, such that different directions were observed for each disease.

ARIMA models were constructed; the parameters, diagnostic plots, and model characteristics are presented in the Supplementary materials (Table S14–17, Fig. S13–20). The observed incidences of gastrointestinal diseases mostly overlapped with the 95% prediction intervals of the predicted values (Fig. 3). In addition, unexpected outbreaks of typhoid and enterohemorrhagic *E. coli* infection occurred, resulting in higher observed incidences than expected (Fig. 3A, B, G, H).

**Fig. 3.**
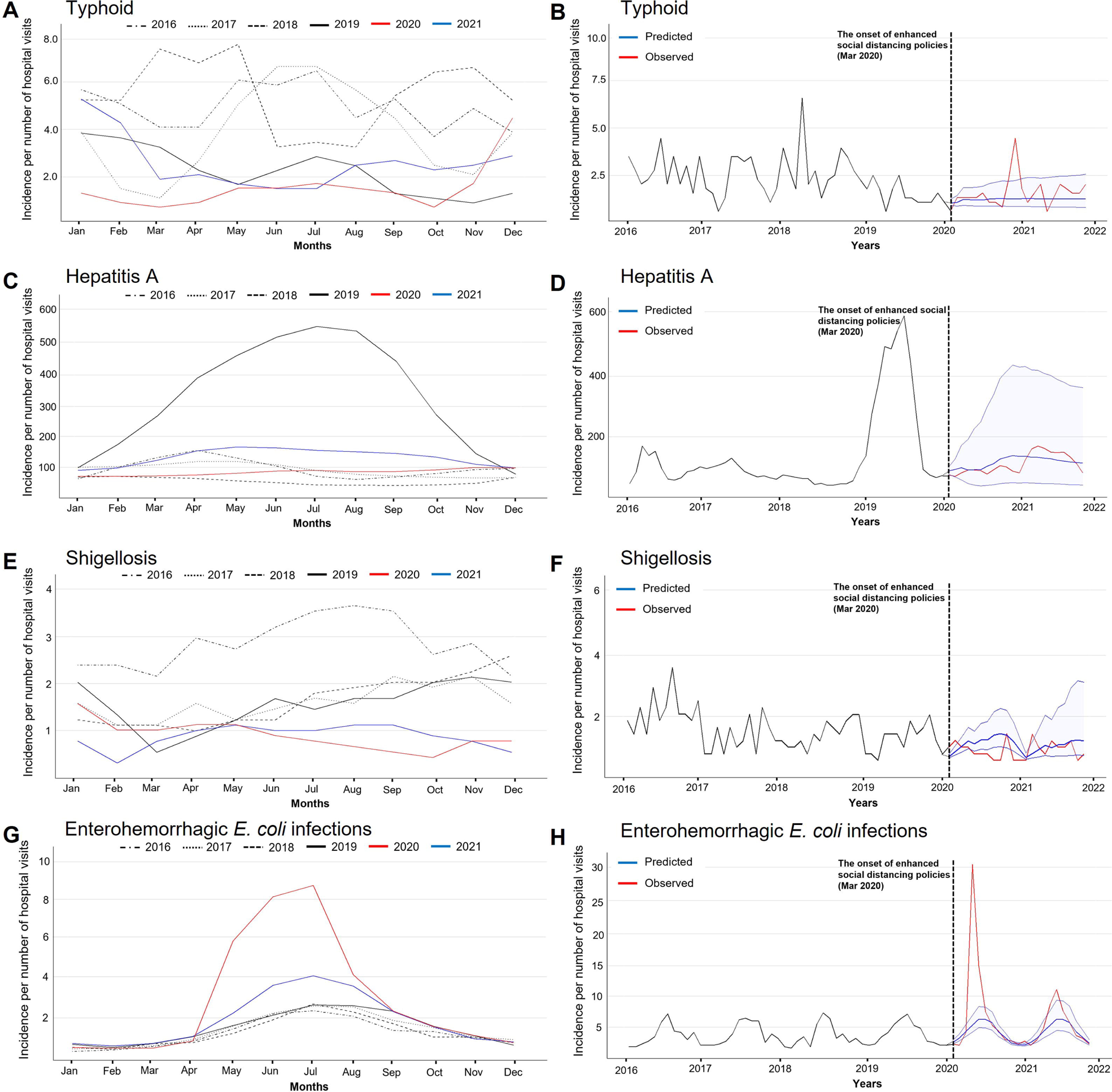
A, C, E, G, Monthly incidences of gastrointestinal infectious diseases from the national surveillance system for notifiable infectious diseases, South Korea, 2016–2019 vs. 2020–2021. B, D, F, H, Observed and predicted monthly incidences from 2016 to 2021.

### 3.3 Incidence of zoonotic diseases in animals

This study included two zoonotic infectious diseases in livestock (Table S5). Comparison of the periods before and after implementation of NPIs showed contrasting patterns in bovine tuberculosis and bovine brucellosis (Table 1). The mean weekly incidences for 2016–2019 per 100,000 cattle varied between diseases: bovine tuberculosis, 2.19; bovine brucellosis, 0.49. Although slight variations were observed among phases, the mean weekly incidences for 2020–2021 after implementation of social distancing measures were as follows: bovine tuberculosis, 1.29; bovine brucellosis, 0.61.

ARIMA models were established; the parameters, diagnostic plots, and model characteristics are presented in the Supplementary materials (Table S18–19, Fig. S21–24). The incidences of bovine tuberculosis were significantly lower than expected from the end of 2020 (Fig. 4A, B). In contrast, the incidence of bovine brucellosis rapidly increased and reached record high case numbers from March 2021 (Fig. 4C, D).

**Fig. 4.**
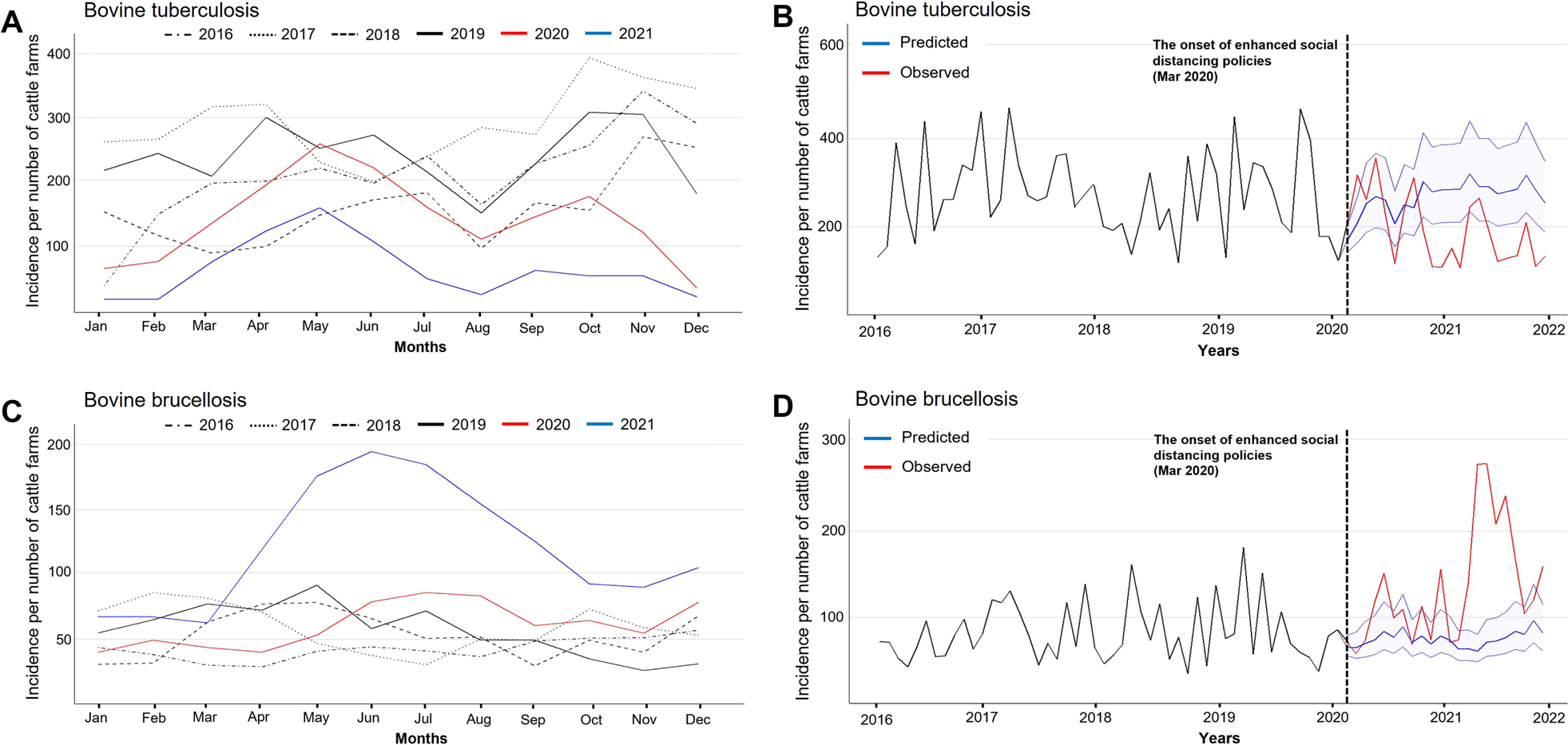
A, C, Monthly incidences of zoonotic infectious diseases in livestock from the national surveillance system, South Korea, 2016–2019 vs. 2020–2021. B, D, Observed and predicted monthly incidences from 2016 to 2021.

## 4. Discussion

The present study utilized national surveillance data regarding notifiable infectious diseases in South Korea from 2016 to 2021 to examine how NPI implementation to control the COVID-19 pandemic affected the patterns of various contagious diseases. We used data from 2016 to 2019 to develop a reliable time series model, then predicted the incidences of communicable diseases for 2020 and 2021 under the assumption that NPIs had not been implemented. By comparing model-predicted values with observed values, we found that the incidence of respiratory infectious diseases significantly decreased after implementation of NPIs. However, there were no significant differences in the occurrence trends of gastrointestinal infectious diseases and livestock diseases between according to NPI implementation. The overall medical expenses from infectious diseases other than COVID-19 decreased by 5.17% in 2020 and 16.14% in 2021, compared with the predicted values (Table 2). The findings provide valuable insights for the implementation of appropriate control measures during future epidemics.

The significant reductions and continuously low incidences of respiratory infectious diseases in South Korea during the COVID-19 pandemic can mainly be attributed to the extensive adoption of NPIs. Regardless of whether the infectious agent is a bacterium (pertussis, scarlet fever, invasive pneumococcal diseases, and tuberculosis) or a virus (varicella and mumps), respiratory infectious diseases transmitted via droplets, fomites, or direct contact generally exhibited a lower incidence after implementation of NPIs; most of these trends persisted until the end of 2021. The sharp decline in respiratory infectious disease incidence after implementation of NPIs was consistent with the findings of previous studies in South Korea regarding the occurrence trends of respiratory infectious diseases [5, 15–17, 39, 40] and the findings of studies focused on respiratory infectious disease patterns in other countries, such as China and the United States [9, 41–43]. After implementation of NPIs, the number of mumps cases remained lower than predicted. However, beginning in October 2021, the number of cases increased above the expected value. This could be attributed to the nationwide relaxation of school attendance criteria in the fall semester of 2021, which led to more outbreaks in schools. Mumps is commonly observed among adolescents aged 13–18 years, and it frequently spreads in settings where people engage in group activities (e.g., schools) [44]. Therefore, precautions are needed to prevent its resurgence after the cessation of NPIs.

Tuberculosis showed a slightly different pattern compared with other respiratory infectious diseases. In the early stages of NPI implementation, the number of cases significantly decreased. However, from the second half of 2020, there was little to no difference between the observed values and the predicted values. Reductions in tuberculosis notifications in early 2020 because of complex factors affecting disease diagnosis have been reported in several countries, including South Korea [19, 45]. However, the impact of NPIs known to prevent acute infections has been limited in suppressing the number of tuberculosis cases in the medium to long term in South Korea, as a significant portion of tuberculosis cases in the country is presumed to result from latent tuberculosis infection progressing to active disease [19, 46]. Thus, there is a need to incorporate the existing strategy of focusing on prophylactic treatment to prevent new infections, as well as the strategy of effectively preventing active tuberculosis onset by treating latent infections.

The incidences of gastrointestinal diseases did not significantly decrease after the implementation of NPIs. Studies using data from countries such as China [9] and the United States [47] showed significant decreases in most gastrointestinal infectious diseases after NPI implementation. Although the dissimilar contexts hinder direct comparisons, differences in the extent of NPI implementation and accessibility to medical services could explain the discrepancies. In the early stages of the COVID-19 pandemic, China and the United States implemented strict social distancing measures and emphasized stay-at-home orders. In contrast, South Korea implemented less strict policies that focused on personal hygiene measures. Therefore, the effectiveness of NPIs in controlling infectious diseases may have varied among countries, and the decrease in health care facility utilization may have been smaller in South Korea [18, 19, 47]. In addition, the gastrointestinal diseases included in this study are primarily classified as foodborne diseases that occur via consumption of contaminated food or water [20]. Therefore, the occurrence of foodborne diseases included in this study might not have been significantly affected by personal hygiene enhancement and social distancing measures.

This study revealed inconsistent temporal trends between the two target zoonotic diseases in industrial animals: bovine tuberculosis and brucellosis. The increased incidence of brucellosis was consistent with prior predictions. Social distancing is likely to hinder proper veterinary care and restrict logistical activity necessary for livestock management [11]. Moreover, in South Korea, the number of cattle farms increased during social distancing, possibly because of the increased cost of beef [48]. The sudden increases in disease incidence may indicate an increased number of inexperienced owners, which would impact management quality. Because the primary route of brucellosis transmission is the movement of infected cattle [49], inexperienced owners may require greater screening skills. In contrast, the decreased incidence of bovine tuberculosis differed from our expectations. One possible explanation is that there was an increased level of bovine tuberculosis surveillance in South Korea. The number of cattle screened for bovine tuberculosis infection has increased since 2017, and the corresponding budget has also increased (Table S20) [50]. Because early detection by effective surveillance plays a key role in controlling chronic diseases with a long latent period, the decreased incidence may be explained by effective surveillance efforts.

This study has several limitations. First, the occurrences of infectious diseases are influenced by various factors, including population immunity, seasonal changes, climatic factors, and human mobility patterns. Thus, it is challenging to make causal inferences regarding the effects of social distancing measures and changes in disease patterns. Therefore, we can only interpret and analyze potential influencing factors. Second, the observed decreases in certain infectious disease incidences may not solely reflect actual reductions in incidence rates. They could also be influenced by other pandemic-related factors including health care utilization. Therefore, we utilized annual hospital visits and health insurance claims to adjust for these changes in health care utilization. However, it is possible that biases persisted with respect to altered health care-seeking behaviors and surveillance capacity. Third, although ARIMA is a well-established and practical technology for infectious disease forecasting [22, 43], it has limitations with respect to distinguishing among transmission factors, such as genetic strains and latent infections. In addition, ARIMA may not be the most appropriate method for long-term predictions, which reduces our confidence in longer-term predictions. Fourth, this study did not consider demographic information, such as age and sex.

## 5. Conclusion

We examined the impacts of COVID-19 and COVID-19-related NPIs on notifiable infectious diseases with diverse transmission pathways throughout the duration of NPI implementation in South Korea. The implementation of NPIs significantly reduced the incidences of infectious diseases transmitted via respiratory routes or direct person-to-person contact; this trend continued until late 2021. Although it is difficult to identify a single factor responsible for changes in the incidences of infectious diseases, the concurrent implementation of NPIs at various levels (individual, community, environmental, and national), along with behavioral changes, likely played a key role in reducing community transmission and alleviating the associated health care burden. Therefore, these comprehensive NPI strategies are important public health considerations for infectious disease control and future pandemic preparedness.

## Data Availability

All data produced in the present study are available upon reasonable request to the authors

## Acknowledgements

This research was supported by the Bio & Medical Technology Development Program of the National Research Foundation, funded by the Korean government (No. 2021M3E5E3081366). However, the funders had no role in the design and conduct of the study; collection, management, analysis, and interpretation of the data; preparation, review, or approval of the paper; or the decision to submit the article for publication.

## CRediT authorship contribution statement

**Taehee Chang**: conceptualization (equal), data curation (lead), formal analysis (lead), funding acquisition (supporting), methodology (equal), visualization (lead) writing – original draft (lead), writing – reviewing and editing (lead); **Sung-il Cho**: conceptualization (supporting), formal analysis (supporting), funding acquisition (supporting), methodology (equal), project administration (equal), writing – reviewing and editing (equal); **Dae sung Yoo**: conceptualization (supporting), formal analysis (supporting), funding acquisition (supporting), methodology (equal), project administration (equal), writing – reviewing and editing (equal); **Kyung-Duk Min**: conceptualization (equal), formal analysis (supporting), funding acquisition (lead), methodology (equal), project administration (equal), visualization (supporting), writing – reviewing and editing (equal)

## Declaration of Competing Interest

The authors declare that they have no known competing financial interests or personal relationships that could have appeared to influence the work reported in this paper.

**Appendix A. Supplementary material**

