## Supplemental materials for "Trends in Nationally Notifiable Infectious Diseases in Humans and Animals During the COVID-19 Pandemic in South Korea"

### Table of Contents

**Supplementary Table 1.** The information on annual hospital visits and annual health insurance claims in Korea……………………………………………………………………………………………….. 3

**Supplementary Table 2.** Three-tier Social Distancing System in Response to Coronavirus Disease 2019 Outbreak, South Korea (June 28, 2020 to November 2020) ……………………………………….. 3

**Supplementary Table 3.** Five-tier Social Distancing System in Response to Coronavirus Disease 2019 Outbreak, South Korea (November 2020 to July 2021) …………………………………………….. 4

**Supplementary Table 4.** Four-tier Social Distancing System in Response to Coronavirus Disease 2019 Outbreak, South Korea July 2021 to onward) ……………………………………………………….. 4

**Supplementary Table 5.** The list of target diseases. ……………………………………………….. 5

**Supplementary Table 6.** Serial intervals of respiratory diseases and their standard deviations.......... 6

**Supplementary Table 7.** Parameters and diagnostic statistics of candidate ARIMA models of Varicella incidences. …………………………………………………………………………………………….. 6

**Supplementary Table 8.** Parameters and diagnostic statistics of candidate ARIMA models of Pertussis incidences. …………………………………………………………………………………………….. 7

**Supplementary Table 9.** Parameters and diagnostic statistics of candidate ARIMA models of Mumps incidences. …………………………………………………………………………………………….. 7

**Supplementary Table 10.** Parameters and diagnostic statistics of candidate ARIMA models of Mumps incidences. …………………………………………………………………………………………….. 8

**Supplementary Table 11.** Parameters and diagnostic statistics of candidate ARIMA models of invasive pneumococcal disease incidences…………………………………………………………………….. 8

**Supplementary Table 12.** Parameters and diagnostic statistics of candidate ARIMA models of scarlet fever incidences. …………………………………………………………………………………….. 9

**Supplementary Table 17.** Parameters and diagnostic statistics of candidate ARIMA models of Enterohemorrhagic *E. coli* incidences. ……………………………………………………………….. 12

**Supplementary Table 18.** Parameters and diagnostic statistics of candidate ARIMA models of Bovine tuberculosis incidences. ………………………………………………………………………………13

**Supplementary Table 19.** Parameters and diagnostic statistics of candidate ARIMA models of Bovine brucellosis incidences. ……………………………………………………………………………… 13

**Supplementary Table 20.** The number of livestock tested for bovine tuberculosis infection and brucellosis and the budget allocated for the testing in the respective operation. ………..……..………. 14

**Supplementary Figure 9.** ACF and PACF plots of scarlet fever incidences………………….……....19

**Supplementary Figure 10.** Residual plots of ARIMA of scarlet fever incidences……….….……....19

**Supplementary Figure 11.** ACF and PACF plots of Tuberculosis incidences…………………..……..20

**Supplementary Figure 12.** Residual plots of ARIMA of Tuberculosis incidences……………………..20

**Supplementary Figure 13.** ACF and PACF plots of Typhoid incidences……………………………… 21

**Supplementary Figure 14.** Residual plots of ARIMA of Typhoid incidences…………………….…. 21

**Supplementary Figure 15.** ACF and PACF plots of Shigellosis incidences…………………….……. 22

**Supplementary Figure 16.** Residual plots of ARIMA of Shigellosis incidences………………….…. 22

**Supplementary Figure 20.** Residual plots of ARIMA of Enterohemorrhagic *E. coli* incidences…….24

**Supplementary Figure 21.** ACF and PACF plots of Bovine tuberculosis incidences………………….25

**Supplementary Figure 22.** Residual plots of ARIMA of Bovine tuberculosis incidences…………….25

**Supplementary Figure 23.** ACF and PACF plots of Bovine brucellosis incidences……………..….26

**Supplementary Figure 24.** Residual plots of ARIMA of Bovine brucellosis incidences…………..….26

**Supplementary Table 1. The information on annual hospital visits and annual health insurance claims in Korea.**

| Year | Annual hospital visits | Annual health insurance claims |
|---|---|---|
| 2010 | 46,690,017 | 1,255,867,542 |
| 2011 | 47,354,032 | 1,280,066,253 |
| 2012 | 47,468,197 | 1,363,271,451 |
| 2013 | 47,756,770 | 1,368,207,434 |
| 2014 | 48,278,590 | 1,393,912,709 |
| 2015 | 48,560,635 | 1,389,266,897 |
| 2016 | 49,011,735 | 1,434,276,197 |
| 2017 | 49,173,457 | 1,452,720,186 |
| 2018 | 49,472,060 | 1,475,254,622 |
| 2019 | 49,634,160 | 1,516,941,079 |
| 2020 | 48,570,460 | 1,321,183,845 |
| 2021 | 49,140,278 | 1,306,717,932 |

**Supplementary Table 2. Three-tier Social Distancing System in Response to Coronavirus Disease 2019 Outbreak, South Korea (June 28, 2020 to November 2020)**

| Social Distancing | Level 1 | Level 2 | Level 3 |
|---|---|---|---|
| The general terms | Distancing in Daily Life | Social Distancing, Moderate Distancing | Intensive Social Distancing |
| The average number of confirmed cases over a period of two weeks (excluding imported cases) | If the average number of confirmed cases is 50 or less | If the number is between 50 and 100 with a regional surge | If the number is 100 or more and doubles at least twice in one week or more |

| | | | |
|---|---|---|---|
| Key Messages | Adherence to hygiene guidelines, allowing for normal economic activities | Avoiding unnecessary outings, gatherings, and use of crowded facilities | Essential social and economic activities should be prioritized, while all other activities should be avoided |

**Supplementary Table 3. Five-tier Social Distancing System in Response to Coronavirus Disease 2019 Outbreak, South Korea (November 2020 to July 2021)**

| Social Distancing | Level 1 | Level 1.5 | Level 2 | Level 2.5 | Level 3 |
|---|---|---|---|---|---|
| The general terms | Distancing in Daily Life | Local Outbreak Initiation | Rapid Local Spread, Nationwide Spread Initiation | Nationwide Outbreak Intensification | Nationwide Major Epidemic |
| Condition of medical system | Routine Management | Localized Threat | Difficulty in Local Response | Exceeding Nationwide Response Capacity | Risk of Healthcare System Collapse |
| Exhibitions, Trade Shows, International Conferences | Consultation with local authorities required with 500 or more participants | Prohibited with 100 or more participants | 1 person per 4m² (approximately 1 pyeong) occupancy limit | 1 person per 16m² (approximately 5 pyeong) occupancy limit | Prohibited if there are 10 or more participants |
| Other Gatherings, Meetings, Events | | | Prohibited with 100 or more participants | Prohibited with 50 or more participants | |

**Supplementary Table 4. Four-tier Social Distancing System in Response to Coronavirus Disease 2019 Outbreak, South Korea July 2021 to onward)**

| Social Distancing | Level 1 | Level 2 | Level 3 | Level 4 |
|---|---|---|---|---|
| The general terms | Sustained Suppression Phase | Regional Outbreak | Regional Epidemic | Nationwide Epidemic |

| | | | | |
|---|---|---|---|---|
| Condition of medical system | Conventional medical response | Limited regional infection control | Limited regional infection control (intensified) | Limited nationwide infection control |
| Overview of Response | Adherence to facility-specific and individual-level hygiene measures to prevent crowding, confined spaces, and close contact. | Restriction on the number of people allowed in a facility or venue. | Prohibition of private gatherings or social events. | Avoidance of unnecessary outings or travel, Encouragement to stay at home and minimize outdoor activities. |
| Criteria: cases per week per 100,000 population | Less than 1 case | 1 or more cases | 2 or more cases | 4 or more cases |

**Supplementary Table 5. The list of target diseases.**

| Disease category | Number | Disease name | Temporal range | Temporal resolution | Modeling outcome |
|---|---|---|---|---|---|
| Respiratory | 1 | Varicella | 2016~2021 | Weekly | Cases (Calculated from Rt) |
| | 2 | Pertussis | 2016~2021 | | |
| | 3 | Mumps | 2016~2021 | | |
| | 4 | Invasive pneumococcal disease | 2016~2021 | | |
| | 5 | Scarlet fever | 2016~2021 | | |
| | 6 | Tuberculosis | 2016~2021 | Monthly | Cases |
| Gastro-intestinal | 1 | Thyphoid fever | 2016~2021 | Monthly | Cases |
| | 2 | Shigellosis | 2016~2021 | | |
| | 3 | Hepatitis A | 2016~2021 | | |
| | 4 | Enterohemorrhagic *Escherichia coli* | 2016~2021 | | |
| Livestock diseases (zoonotic) | 1 | Cattle tuberculosis | 2016~2021 | Monthly | Cases |

| | | |
|---|---|---|
| 2 | Cattle brucellosis | 2016~2021 |

**Supplementary Table 6. Serial intervals of respiratory diseases and their standard deviations.**

| Respiratory disease | Serial interval (days) | Standard deviation (days) |
|---|---|---|
| Varicella | 14.0 | 2.2 |
| Pertussis | 22.8 | 6.5 |
| Mumps | 18.0 | 3.5 |
| Invasive pneumococcal diseases (Streptococcus pneumoniae) | 6.6 | 1.8 |
| Scarlet fever | 14.0 | 4.9 |

**Supplementary Table 7. Parameters and diagnostic statistics of candidate ARIMA models of Varicella incidences.**

| **Model** | **AICc** | **AIC** | **BIC** | **RMSE** |
|---|---|---|---|---|
| ARIMA(3,0,2)(0,1,0)[52] | -618.75 | -619.34 | -601.23 | 0.03 |
| ARIMA(3,0,1)(0,1,0)[52] | -568.56 | -568.98 | -553.89 | 0.03 |
| ARIMA(2,0,2)(0,1,0)[52] | -567.91 | -568.32 | -553.23 | 0.04 |
| ARIMA(3,1,2)(0,1,0)[52] | -616.92 | -617.51 | -599.44 | 0.03 |
| ARIMA(3,0,2)(1,1,0)[52] | -607.72 | -608.5 | -587.38 | 0.03 |
| ARIMA(3,0,2)(0,1,1)[52] | -591.79 | -592.57 | -571.45 | 0.03 |
| ARIMA(4,0,2)(0,1,0)[52] | -586.47 | -587.25 | -566.13 | 0.04 |
| ARIMA(3,0,3)(0,1,0)[52] | -579.71 | -580.49 | -559.37 | 0.04 |
| ARIMA(4,0,3)(0,1,0)[52] | -588.23 | -589.24 | -565.11 | 0.04 |
| ARIMA(4,1,3)(0,1,0)[52] | -612.51 | -613.53 | -589.44 | 0.03 |

**Supplementary Table 8. Parameters and diagnostic statistics of candidate ARIMA models of Pertussis incidences.**

| Model | AICc | AIC | BIC | RMSE |
|---|---|---|---|---|
| ARIMA(4,1,0)(1,1,0)[52] | -560.81 | -561.45 | -543.80 | 0.05 |
| ARIMA(4,1,0)(1,0,0)[52] | -868.08 | -868.53 | -848.99 | 0.03 |
| ARIMA(4,1,0)(0,0,1)[52] | -868.14 | -868.60 | -849.05 | 0.03 |
| ARIMA(4,1,0)(1,0,1)[52] | -868.15 | -868.76 | -845.96 | 0.03 |
| ARIMA(4,1,1)(1,1,1)[52] | -866.38 | -867.17 | -841.11 | 0.03 |
| ARIMA(5,1,0)(1,0,1)[52] | -867.09 | -867.88 | -841.82 | 0.03 |
| ARIMA(3,1,0)(1,0,1)[52] | -852.7 | -853.15 | -833.61 | 0.04 |
| ARIMA(4,1,2)(1,0,1)[52] | -877.79 | -878.77 | -849.46 | 0.04 |
| ARIMA(4,0,3)(0,1,0)[52] | -588.23 | -589.24 | -565.11 | 0.03 |
| ARIMA(4,1,3)(0,1,0)[52] | -612.51 | -613.53 | -589.44 | 0.03 |

**Supplementary Table 9. Parameters and diagnostic statistics of candidate ARIMA models of Mumps incidences.**

| Model | AICc | AIC | BIC | RMSE |
|---|---|---|---|---|
| ARIMA(1,2,2)(1,1,0)[52] | -702.98 | -703.41 | -688.57 | 0.03 |
| ARIMA(1,0,2)(1,1,0)[52] | -788.12 | -788.55 | -773.63 | 0.02 |
| ARIMA(1,0,2)(1,1,1)[52] | -786.45 | -787.06 | -769.15 | 0.02 |
| ARIMA(2,0,2)(1,1,0)[52] | -774.36 | -774.96 | -757.06 | 0.03 |
| ARIMA(1,0,1)(1,1,0)[52] | -766.38 | -767.17 | -741.11 | 0.02 |
| ARIMA(1,0,1)(0,1,0)[52] | -643.13 | -643.3 | -634.35 | 0.03 |

| | | | | |
|---|---|---|---|---|
| ARIMA(2,0,1)(1,1,0)[52] | -711.38 | -711.81 | -696.89 | 0.04 |
| ARIMA(0,0,1)(1,1,0)[52] | -823.79 | -823.32 | -814.46 | 0.02 |
| ARIMA(1,1,1)(1,1,0)[52] | -709.23 | -689.24 | -665.11 | 0.03 |
| ARIMA(2,1,1)(1,1,0)[52] | -784.51 | -785.53 | -770.44 | 0.03 |

**Supplementary Table 10. Parameters and diagnostic statistics of candidate ARIMA models of Mumps incidences.**

| Model | AICc | AIC | BIC | RMSE |
|---|---|---|---|---|
| ARIMA(1,2,2)(1,1,0)[52] | -702.98 | -703.41 | -688.57 | 0.03 |
| ARIMA(1,0,2)(1,1,0)[52] | -788.12 | -788.55 | -773.63 | 0.02 |
| ARIMA(1,0,2)(1,1,1)[52] | -786.45 | -787.06 | -769.15 | 0.02 |
| ARIMA(2,0,2)(1,1,0)[52] | -774.36 | -774.96 | -757.06 | 0.03 |
| ARIMA(1,0,1)(1,1,0)[52] | -766.38 | -767.17 | -741.11 | 0.02 |
| ARIMA(1,0,1)(0,1,0)[52] | -643.13 | -643.3 | -634.35 | 0.03 |
| ARIMA(2,0,1)(1,1,0)[52] | -711.38 | -711.81 | -696.89 | 0.04 |
| ARIMA(0,0,1)(1,1,0)[52] | -823.79 | -823.32 | -814.46 | 0.02 |
| ARIMA(1,1,1)(1,1,0)[52] | -709.23 | -689.24 | -665.11 | 0.03 |
| ARIMA(2,1,1)(1,1,0)[52] | -784.51 | -785.53 | -770.44 | 0.03 |

**Supplementary Table 11. Parameters and diagnostic statistics of candidate ARIMA models of invasive pneumococcal disease incidences.**

| Model | AICc | AIC | BIC | RMSE |
|---|---|---|---|---|
| ARIMA(2,0,0)(1,1,1)[52] | -570.40 | -570.97 | -552.67 | 0.04 |

| | | | | |
|---|---|---|---|---|
| ARIMA(2,0,1)(1,1,1)[52] | -567.45 | -568.01 | -549.71 | 0.04 |
| ARIMA(1,0,0)(1,1,1)[52] | -334.94 | -335.2 | -323.00 | 0.05 |
| ARIMA(1,0,1)(1,1,1)[52] | -428.78 | -429.18 | -413.93 | 0.04 |
| ARIMA(3,0,0)(1,1,1)[52] | -567.59 | -568.16 | -549.86 | 0.04 |
| ARIMA(3,0,1)(1,1,1)[52] | -570.55 | -571.31 | -559.96 | 0.03 |
| ARIMA(2,0,0)(1,1,0)[52] | -570.50 | -570.76 | -558.56 | 0.04 |
| ARIMA(2,0,0)(0,1,0)[52] | -507.47 | -507.63 | -498.48 | 0.04 |
| ARIMA(2,0,0)(0,1,1)[52] | -557.43 | -557.7 | -545.5 | 0.04 |
| ARIMA(2,0,0)(1,0,1)[52] | -484.51 | -485.53 | -470.44 | 0.03 |

**Supplementary Table 12. Parameters and diagnostic statistics of candidate ARIMA models of scarlet fever incidences.**

| Model | AICc | AIC | BIC | RMSE |
|---|---|---|---|---|
| ARIMA(2,1,3)(0,1,0)[52] | -954.93 | -955.53 | -937.55 | 0.01 |
| ARIMA(2,1,2)(0,1,0)[52] | -952.57 | -953.00 | -938.71 | 0.01 |
| ARIMA(3,1,2)(0,1,0)[52] | -926.83 | -927.42 | -909.44 | 0.01 |
| ARIMA(1,1,2)(0,1,0)[52] | -898.41 | -898.69 | -886.70 | 0.01 |
| ARIMA(2,1,1)(0,1,0)[52] | -946.91 | -947.19 | -935.20 | 0.01 |
| ARIMA(2,1,2)(1,1,0)[52] | -948.49 | -949.09 | -931.11 | 0.01 |
| ARIMA(2,1,2)(1,1,1)[52] | -947.69 | -948.49 | -947.51 | 0.01 |
| ARIMA(2,1,2)(2,1,0)[52] | -855.98 | -856.78 | -835.80 | 0.01 |
| ARIMA(2,1,2)(0,1,1)[52] | -956.58 | -957.17 | -939.19 | 0.01 |
| ARIMA(2,1,2)(2,1,1)[52] | -898.62 | -899.65 | -875.67 | 0.01 |

**Supplementary Table 13. Parameters and diagnostic statistics of candidate ARIMA models of Tuberculosis incidences.**

| Model | AICc | AIC | BIC | RMSE |
|---|---|---|---|---|
| ARIMA(0,1,3)(1,0,0)[12] | -158.37 | -159.76 | -150.32 | 21.87 |
| ARIMA(1,1,3)(1,0,0)[12] | -157.84 | -159.84 | -148.49 | 21.27 |
| ARIMA(1,1,4)(1,0,0)[12] | -157.34 | -160.07 | -146.83 | 19.25 |
| ARIMA(0,1,4)(1,0,0)[12] | -159.54 | -161.54 | -150.19 | 20.21 |
| ARIMA(0,1,4)(1,0,1)[12] | -156.94 | -159.68 | -146.43 | 20.05 |
| ARIMA(0,1,4)(2,0,0)[12] | -156.87 | -159.61 | -146.36 | 20.13 |
| ARIMA(0,1,4)(2,0,1)[12] | -154.22 | -157.82 | -146.53 | 19.92 |
| ARIMA(0,1,5)(1,0,0)[12] | -157.04 | -159.78 | -146.53 | 20.19 |
| ARIMA(0,1,2)(1,0,0)[12] | -149.13 | -150.04 | -142.48 | 24.78 |
| ARIMA(1,1,2)(1,0,0)[12] | -149.96 | -151.35 | -141.89 | 24.01 |

**Supplementary Table 14. Parameters and diagnostic statistics of candidate ARIMA models of Typhoid incidences.**

| Model | AICc | AIC | BIC | RMSE |
|---|---|---|---|---|
| ARIMA(0,1,3)(1,0,0)[12] | 22.75 | 21.35 | 30.81 | 0.35 |
| ARIMA(0,1,2)(1,0,0)[12] | 33.85 | 32.94 | 40.51 | 0.42 |
| ARIMA(0,1,4)(1,0,0)[12] | 25.05 | 23.05 | 34.41 | 0.35 |
| ARIMA(0,1,1)(1,0,0)[12] | 31.54 | 31.01 | 36.68 | 0.42 |
| ARIMA(1,1,3)(1,0,0)[12] | 25.19 | 23.19 | 34.54 | 0.35 |
| ARIMA(2,1,3)(1,0,0)[12] | 24.31 | 21.57 | 34.82 | 0.34 |

| | | | | |
|---|---|---|---|---|
| ARIMA(0,1,3)(1,1,0)[12] | 40.07 | 38.13 | 46.19 | 0.42 |
| ARIMA(0,1,3)(1,1,1)[12] | 40.06 | 37.26 | 46.93 | 0.31 |
| ARIMA(0,1,3)(2,0,0)[12] | 25.28 | 23.28 | 34.64 | 0.35 |
| ARIMA(0,1,3)(2,0,1)[12] | 27.49 | 24.75 | 38.01 | 0.34 |

**Supplementary Table 15. Parameters and diagnostic statistics of candidate ARIMA models of Shigellosis incidences.**

| Model | AICc | AIC | BIC | RMSE |
|---|---|---|---|---|
| ARIMA(3,1,0)(0,1,0)[12] | 32.92 | 31.67 | 38.11 | 0.17 |
| ARIMA(3,1,1)(0,1,0)[12] | 34.88 | 32.94 | 41.02 | 0.16 |
| ARIMA(3,1,2)(0,1,0)[12] | 37.28 | 34.48 | 44.15 | 0.16 |
| ARIMA(2,1,0)(0,1,0)[12] | 39.55 | 38.83 | 43.66 | 0.19 |
| ARIMA(1,1,0)(0,1,0)[12] | 38.36 | 38.01 | 41.23 | 0.19 |
| ARIMA(3,1,0)(1,1,0)[12] | 33.58 | 31.65 | 39.77 | 0.16 |
| ARIMA(3,1,0)(0,1,1)[12] | 33.54 | 31.56 | 39.62 | 0.16 |
| ARIMA(3,1,0)(1,1,1)[12] | 36.35 | 33.55 | 43.22 | 0.16 |
| ARIMA(4,1,0)(0,1,0)[12] | 34.73 | 32.88 | 40.85 | 0.16 |
| ARIMA(4,1,1)(0,1,0)[12] | 37.6 | 34.8 | 44.46 | 0.16 |

**Supplementary Table 16. Parameters and diagnostic statistics of candidate ARIMA models of Hepatitis A incidences.**

| Model | AICc | AIC | BIC | RMSE |
|---|---|---|---|---|
| ARIMA(4,1,0) | -53.46 | -54.86 | -45.44 | 18.78 |

| | | | | |
|---|---|---|---|---|
| ARIMA(4,1,1) | -51.17 | -53.17 | -41.82 | 18.54 |
| ARIMA(4,1,2) | -49.74 | -52.48 | -39.23 | 17.74 |
| ARIMA(5,1,0) | -51.77 | -53.77 | -42.42 | 17.83 |
| ARIMA(4,2,0) | -45.73 | -47.16 | -37.82 | 22.49 |
| ARIMA(3,1,0) | -52.91 | -53.82 | -46.25 | 19.75 |
| ARIMA(3,1,1) | -53.13 | -54.53 | -45.07 | 18.85 |
| ARIMA(3,1,2) | -52.08 | -54.08 | -42.73 | 17.47 |
| ARIMA(4,0,0) | -48.93 | -50.88 | -39.41 | 20.56 |
| ARIMA(5,1,1) | -52.33 | -55.06 | -41.82 | 15.99 |

**Supplementary Table 17. Parameters and diagnostic statistics of candidate ARIMA models of Enterohemorrhagic *E. coli* incidences.**

| Model | AICc | AIC | BIC | RMSE |
|---|---|---|---|---|
| ARIMA(1,0,0)(0,1,0)[12] | 2.47 | 2.13 | 5.45 | 0.32 |
| ARIMA(1,0,1)(0,1,0)[12] | 4.62 | 3.92 | 8.83 | 0.32 |
| ARIMA(1,1,1)(0,1,0)[12] | 6.67 | 5.95 | 10.78 | 0.33 |
| ARIMA(1,0,0)(1,1,0)[12] | 3.1 | 2.39 | 7.32 | 0.29 |
| ARIMA(1,0,1)(1,1,0)[12] | 2.82 | 1.61 | 8.15 | 0.24 |
| ARIMA(0,0,0)(0,1,0)[12] | 9.07 | 8.96 | 10.63 | 0.38 |
| ARIMA(2,0,0)(0,1,0)[12] | 4.57 | 3.87 | 8.78 | 0.16 |
| ARIMA(2,0,1)(0,1,0)[12] | 6.34 | 5.13 | 11.68 | 0.31 |
| ARIMA(1,0,1)(1,1,1)[12] | 9.13 | 7.13 | 15.31 | 0.31 |
| ARIMA(1,0,0)(0,2,0)[12] | 28.73 | 97.28 | 31.22 | 0.47 |

**Supplementary Table 18. Parameters and diagnostic statistics of candidate ARIMA models of Bovine tuberculosis incidences.**

| Model | AICc | AIC | BIC | RMSE |
|---|---|---|---|---|
| ARIMA(0,0,2) | -25.6 | -26.49 | 18.84 | 40.37 |
| ARIMA(1,0,2)(1,0,0)[12] | -21.3 | -23.26 | -11.78 | 40.49 |
| ARIMA(1,0,2)(1,0,1)[12] | -20.36 | -23.03 | -9.64 | 37.47 |
| ARIMA(1,0,2)(2,0,1)[12] | -21.02 | -24.53 | -9.24 | 35.02 |
| ARIMA(1,0,3)(2,0,1)[12] | -18.05 | -22.55 | -5.34 | 34.54 |
| ARIMA(1,0,3)(2,0,2)[12] | -15.77 | -21.41 | -2.29 | 33.64 |
| ARIMA(2,0,2)(2,0,1)[12] | -18.15 | -22.65 | -5.44 | 33.19 |
| ARIMA(2,0,2)(2,0,2)[12] | -15.95 | -21.6 | -2.48 | 31.56 |
| ARIMA(2,0,2)(2,0,3)[12] | -11.66 | -18.61 | 2.42 | 37.07 |
| ARIMA(2,0,1)(2,0,3)[12] | -4.24 | -9.88 | 9.24 | 36.74 |

**Supplementary Table 19. Parameters and diagnostic statistics of candidate ARIMA models of Bovine brucellosis incidences.**

| Model | AICc | AIC | BIC | RMSE |
|---|---|---|---|---|
| ARIMA(0,0,2) | 17.08 | 16.19 | 23.84 | 12.46 |
| ARIMA(1,0,2)(1,0,0)[12] | 20.10 | 18.14 | 29.62 | 11.97 |
| ARIMA(1,0,2)(2,0,1)[12] | 22.44 | 18.93 | 34.23 | 11.06 |
| ARIMA(2,0,2)(2,0,1)[12] | 19.72 | 24.22 | 36.93 | 10.89 |
| ARIMA(1,0,3)(2,0,1)[12] | 20.11 | 24.61 | 37.32 | 11.23 |
| ARIMA(1,0,3)(2,0,2)[12] | 20.7 | 26.34 | 39.82 | 9.34 |

| | | | | |
|---|---|---|---|---|
| ARIMA(2,0,2)(2,0,1)[12] | 19.72 | 24.22 | 36.93 | 10.89 |
| ARIMA(2,0,2)(2,0,2)[12] | 20.93 | 26.57 | 40.05 | 9.23 |
| ARIMA(2,0,2)(2,0,3)[12] | 29.71 | 22.76 | 43.8 | 9.01 |
| ARIMA(2,0,1)(2,0,3)[12] | 42.16 | 47.8 | 61.28 | 13.14 |

**Supplementary Table 20. The number of livestock tested for bovine tuberculosis infection and brucellosis and the budget allocated for the testing in the respective operation.**

| **Number of cattle (1,000 numbers)** | **2017** | **2018** | **2019** | **2020** | **2021** |
|---|---|---|---|---|---|
| Dairy cow (PPD*) | 343 | 342 | 340 | 344 | 340 |
| Beef cattle (Certificate of examination) | 462 | 547 | 580 | 625 | 718 |
| Bovine brucellosis (MRT†) | 80 | 77 | 77 | 75 | 75 |
| Bovine brucellosis (RB‡) | 1,938 | 1,891 | 1,858 | 1,824 | 1,824 |
| **Allocated budget (1 USD)** | **2017** | **2018** | **2019** | **2020** | **2021** |
| Dairy cow (PPD) | 239,141 | 238,165 | 236,771 | 239,768 | 253,981 |
| Beef cattle (Certificate of examination) | 1,710,510 | 2,022,790 | 2,146,000 | 2,311,020 | 2,697,240 |
| Bovine brucellosis (MRT) | 26,207 | 25,223 | 25,092 | 24,600 | 24,600 |
| Bovine brucellosis (RB) | 654,639 | 638,931 | 628,173 | 616,953 | 616,953 |

*Purified protein derivatives
†Milk ring test
‡Rose-bengal test

**Supplementary Figure 1. ACF and PACF plots of Varicella incidences**

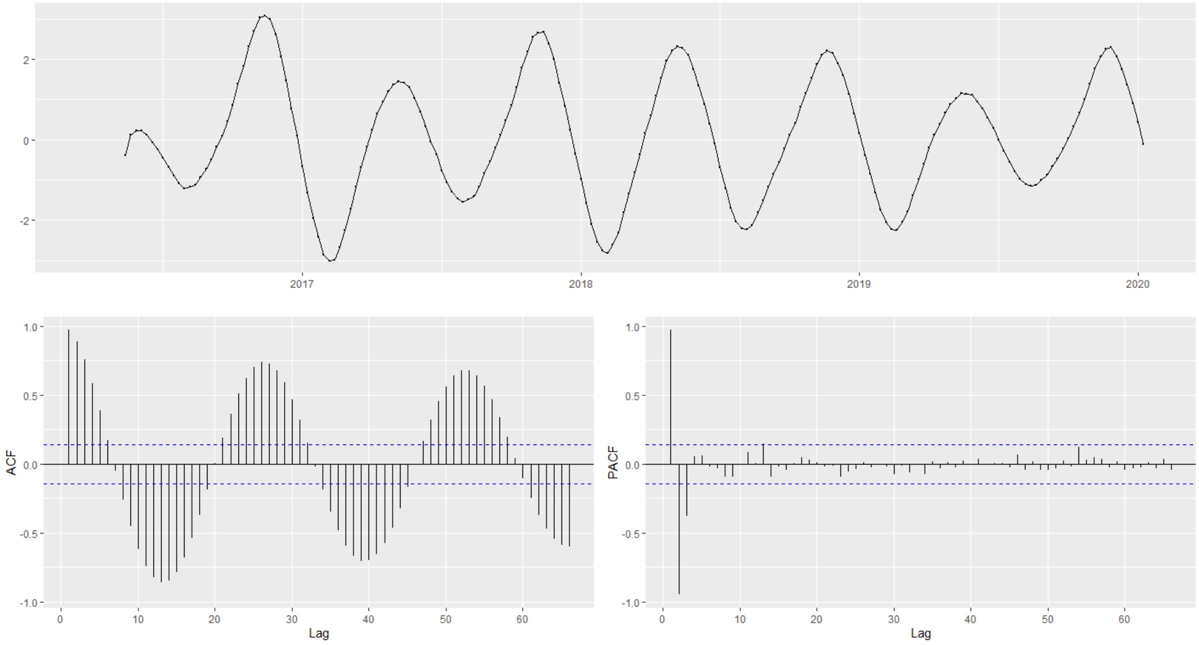

**Supplementary Figure 2. Residual plots of ARIMA of Varicella incidences**

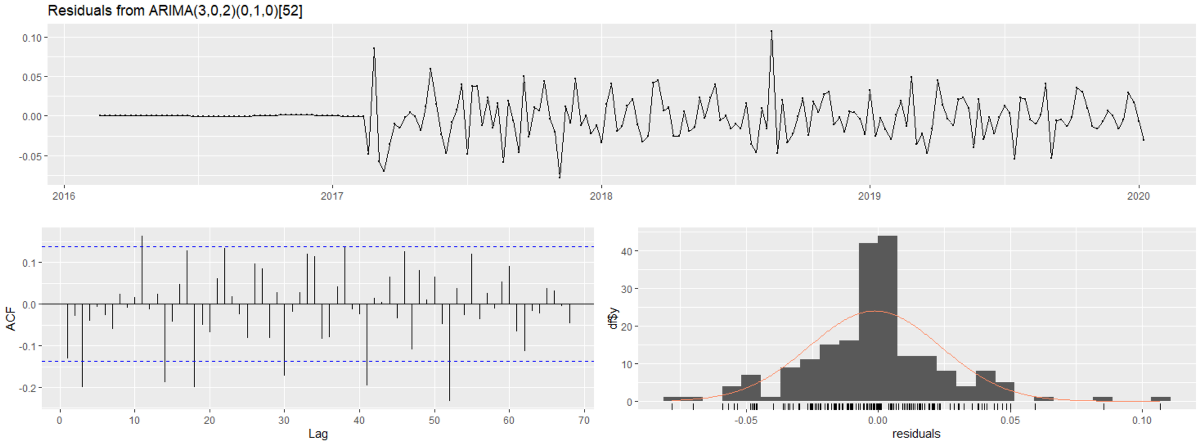

**Supplementary Figure 3. ACF and PACF plots of Pertussis incidences**

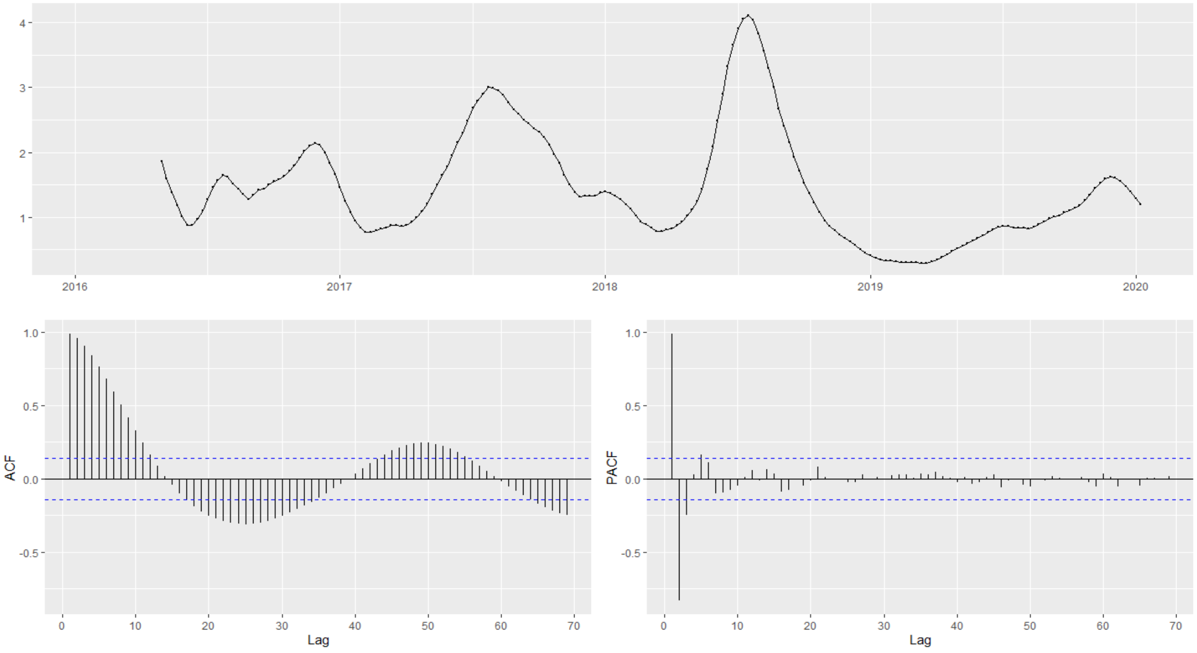

**Supplementary Figure 4. Residual plots of ARIMA of Pertussis incidences**

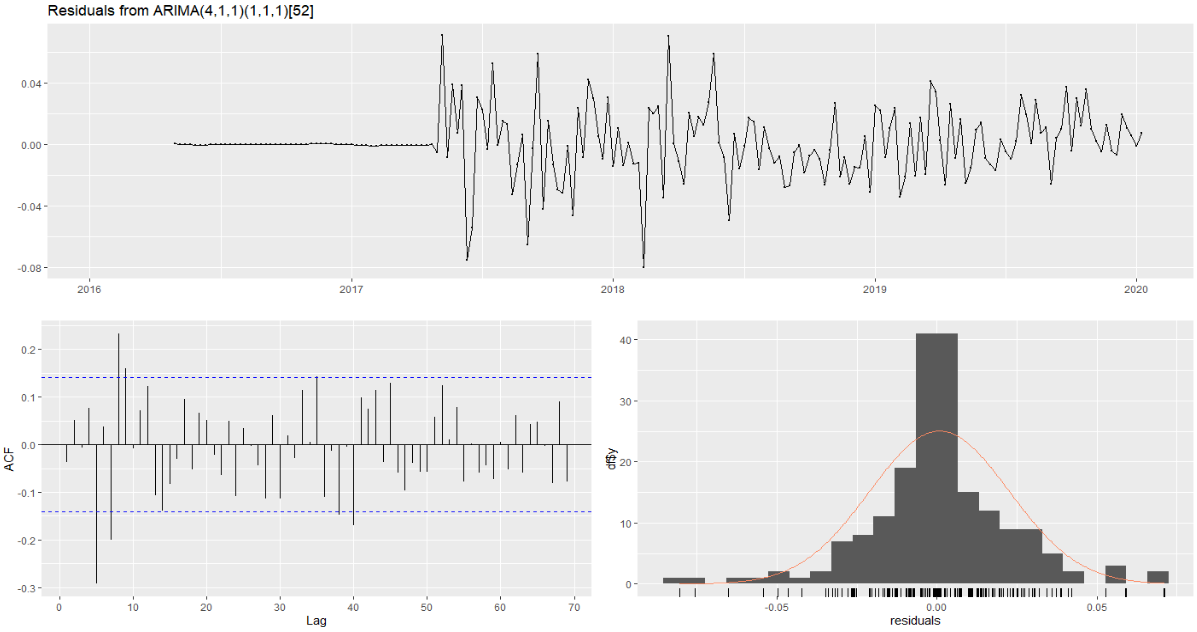

### Supplementary Figure 5. ACF and PACF plots of Mumps incidences

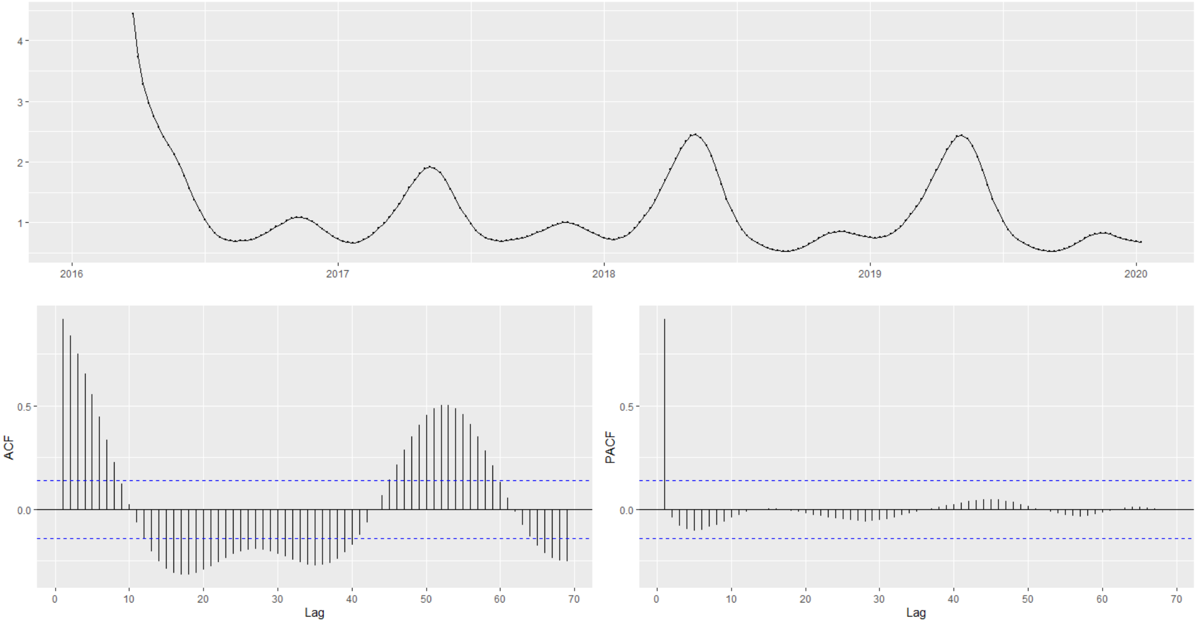

### Supplementary Figure 6. Residual plots of ARIMA of Mumps incidences

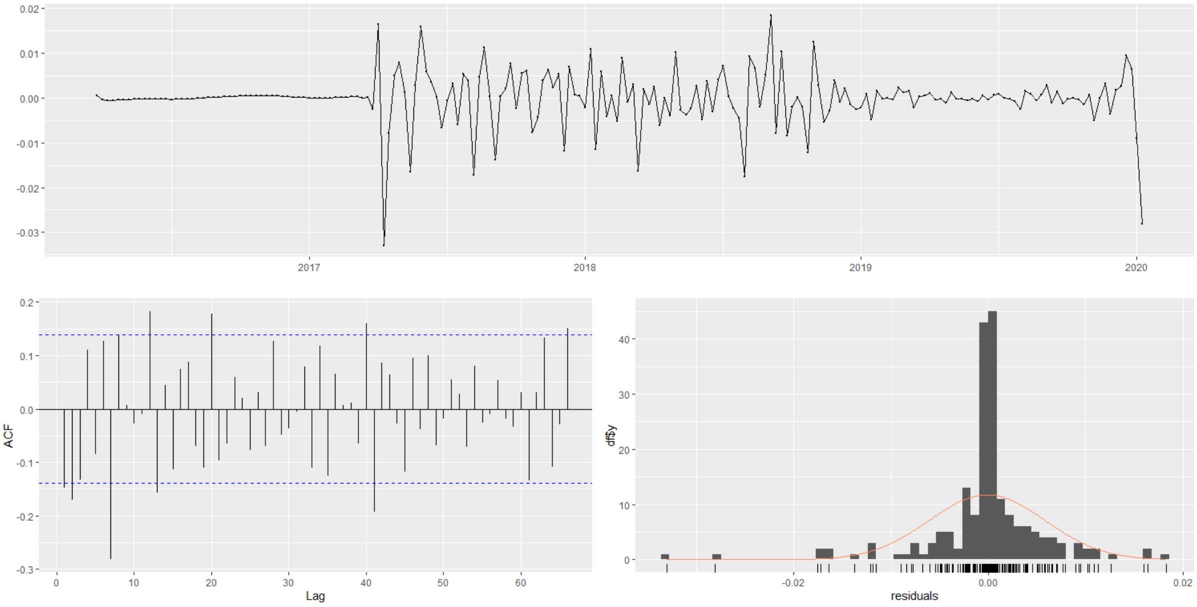

### Supplementary Figure 7. ACF and PACF plots of invasive pneumococcal disease incidences

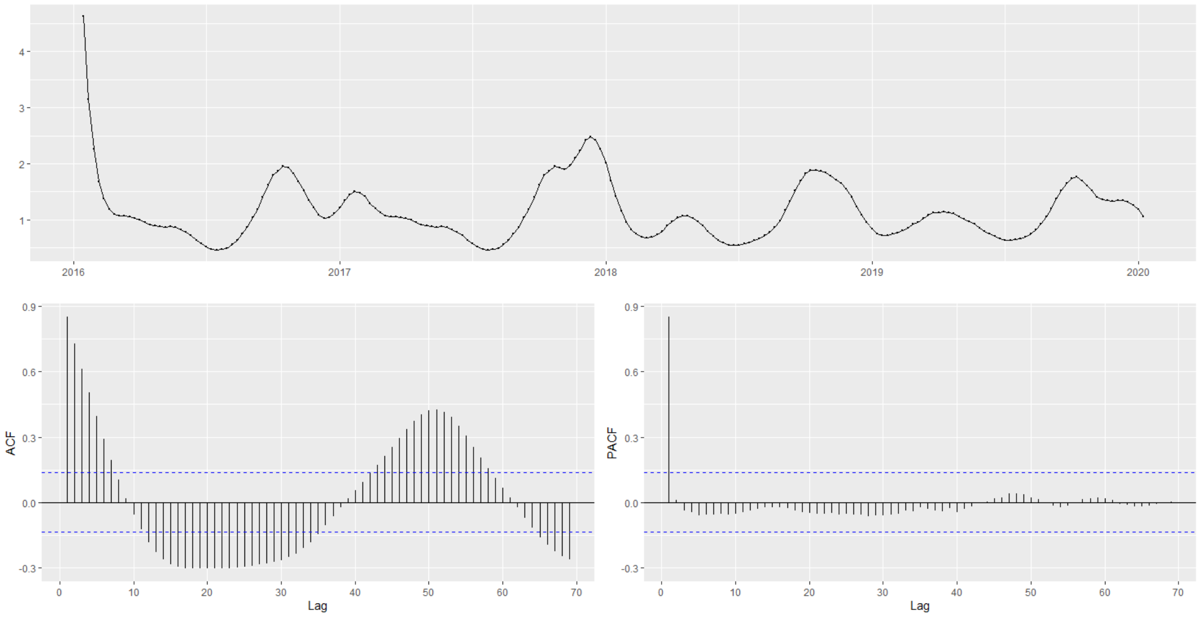

### Supplementary Figure 8. Residual plots of ARIMA of invasive pneumococcal disease incidences

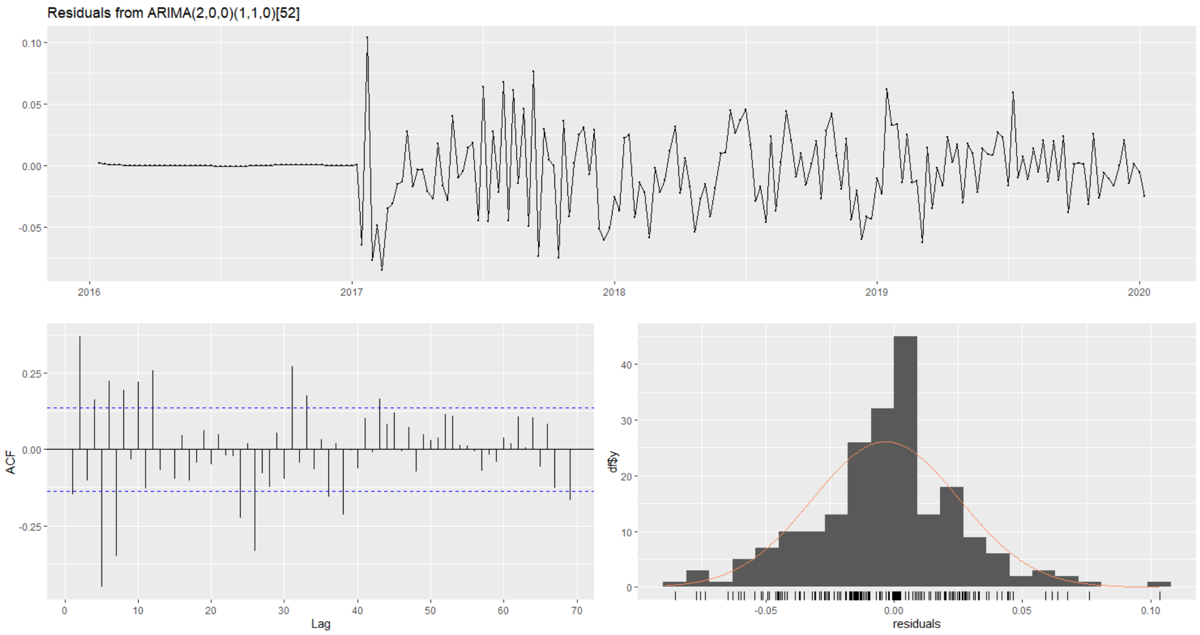

**Supplementary Figure 9. ACF and PACF plots of scarlet fever incidences**

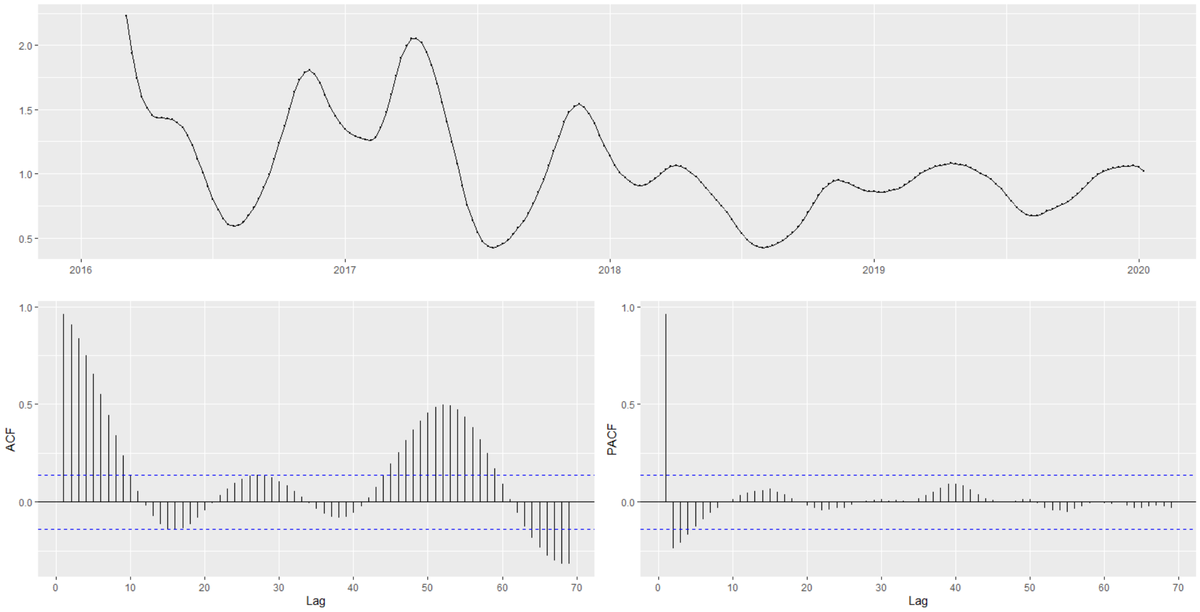

**Supplementary Figure 10. Residual plots of ARIMA of scarlet fever incidences**

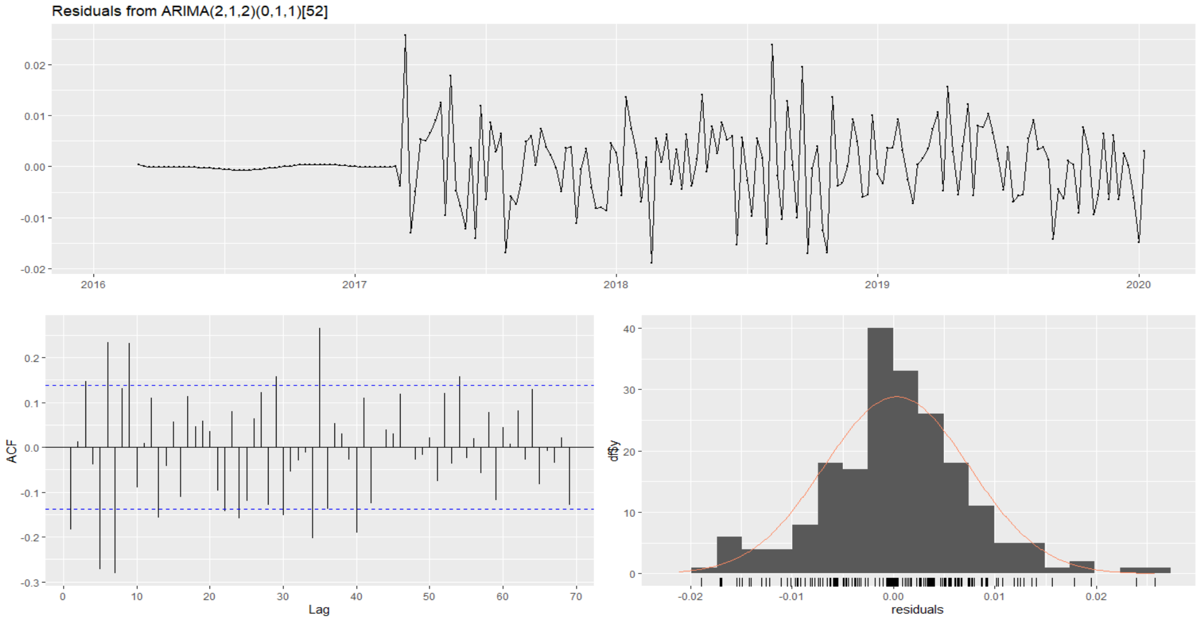

### Supplementary Figure 11. ACF and PACF plots of Tuberculosis incidences

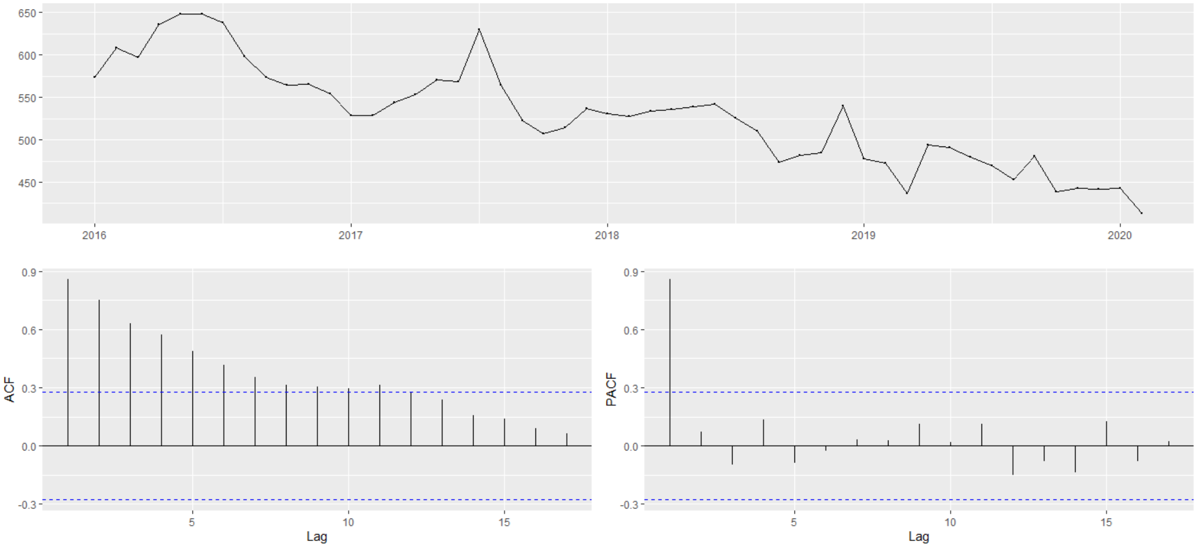

### Supplementary Figure 12. Residual plots of ARIMA of Tuberculosis incidences

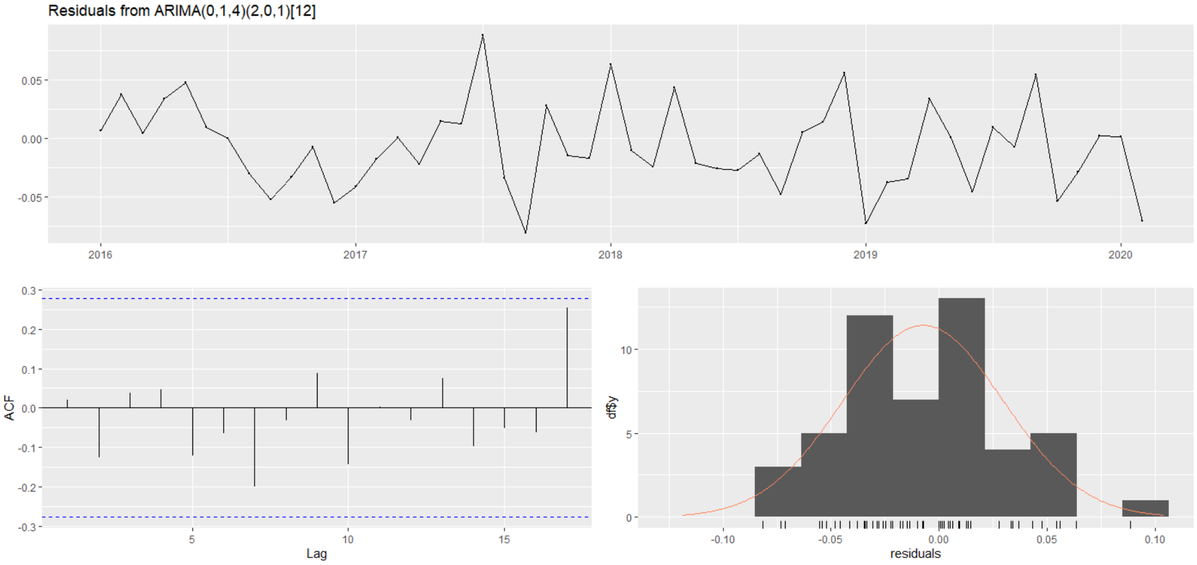

**Supplementary Figure 13. ACF and PACF plots of Typhoid incidences**

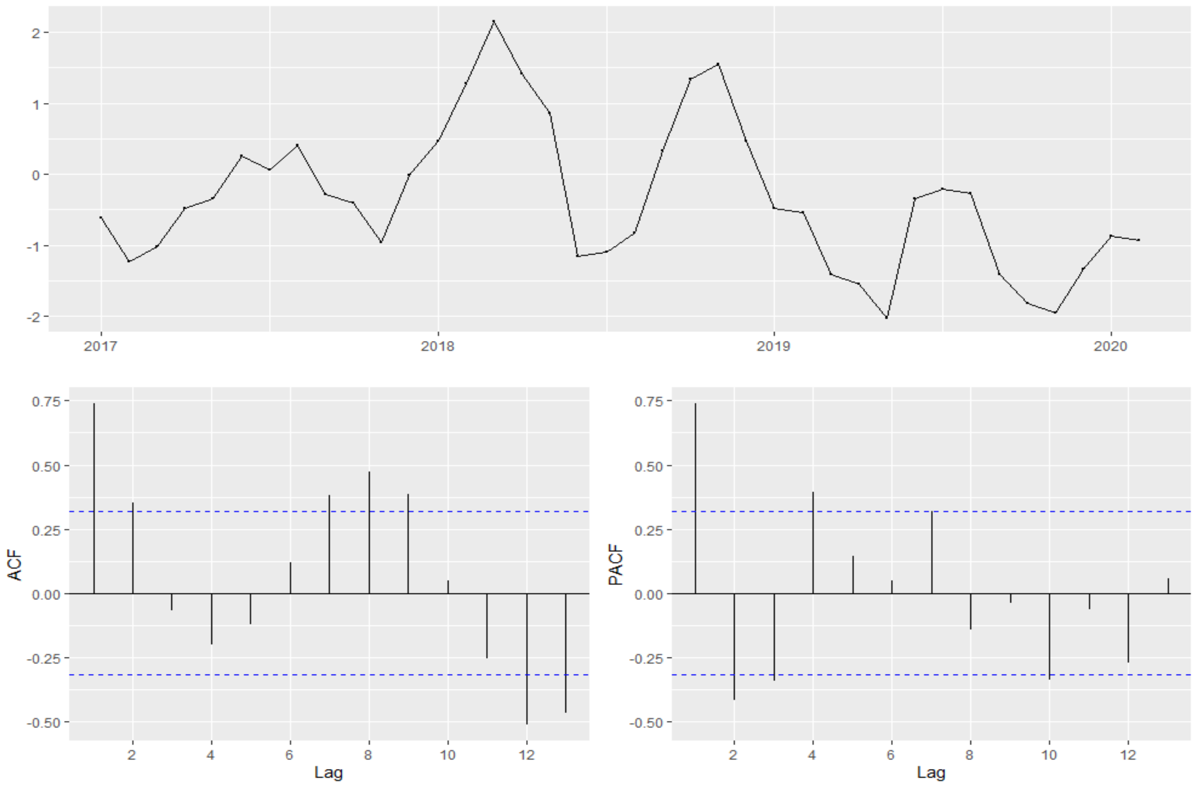

**Supplementary Figure 14. Residual plots of ARIMA of Typhoid incidences**

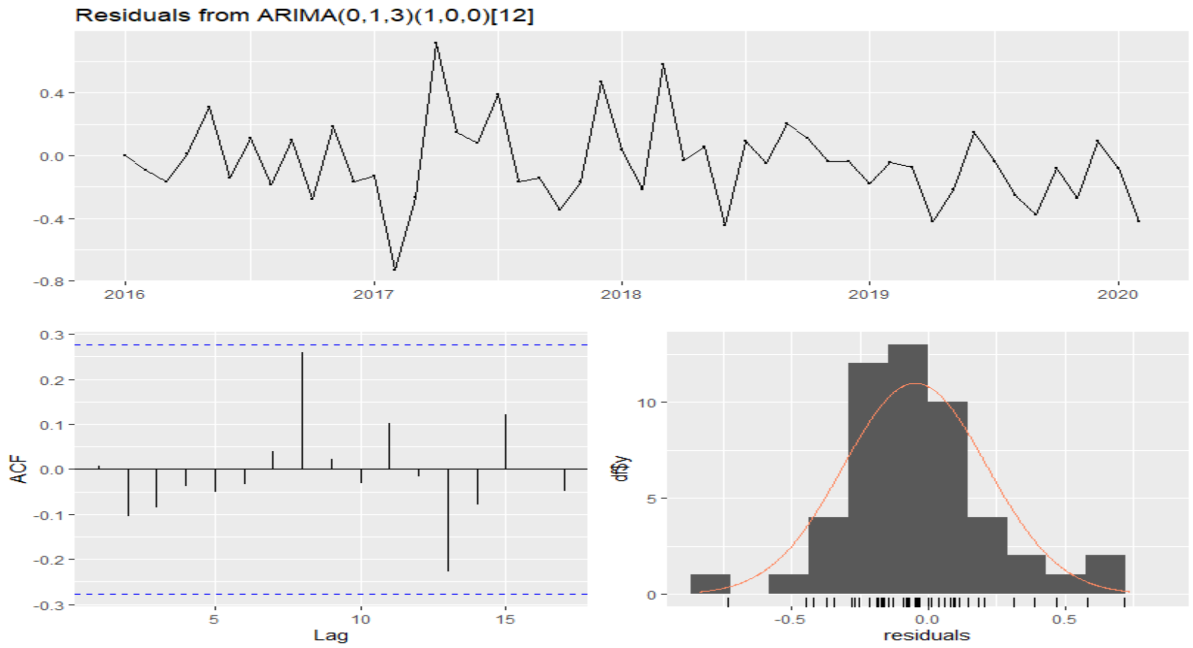

**Supplementary Figure 15. ACF and PACF plots of Shigellosis incidences**

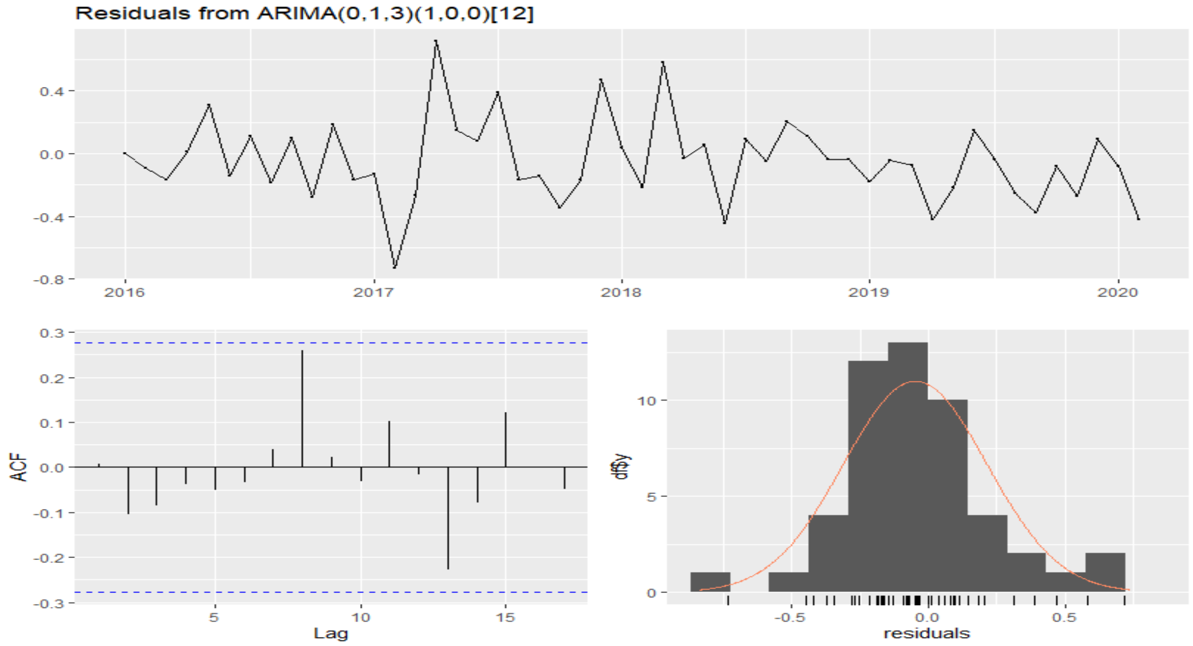

**Supplementary Figure 16. Residual plots of ARIMA of Shigellosis incidences**

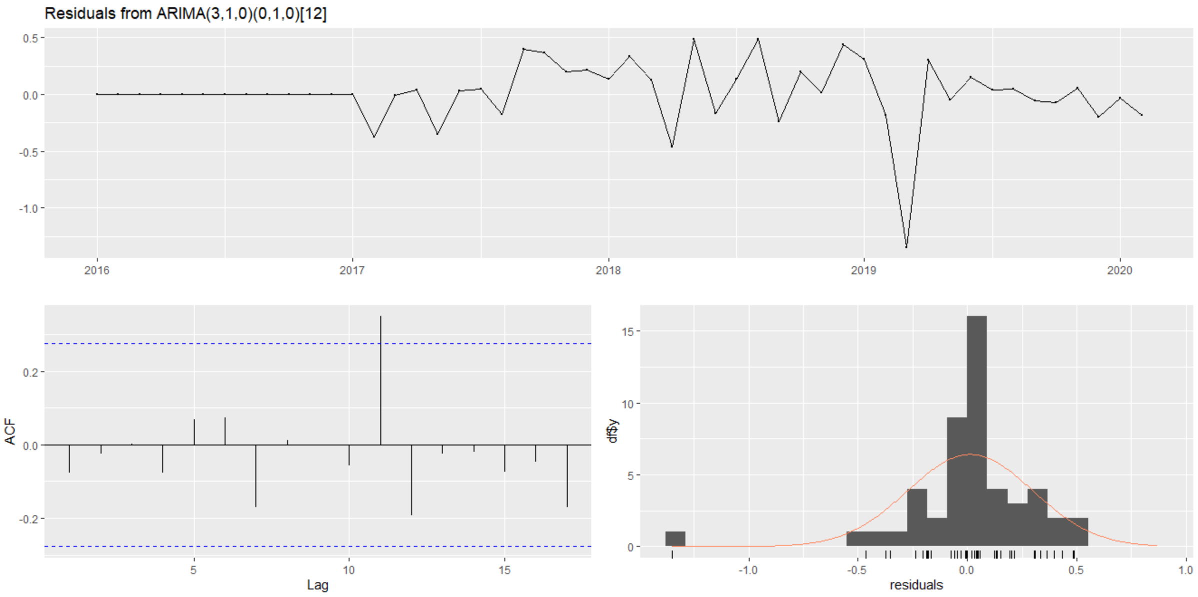

**Supplementary Figure 17. ACF and PACF plots of Hepatitis A incidences**

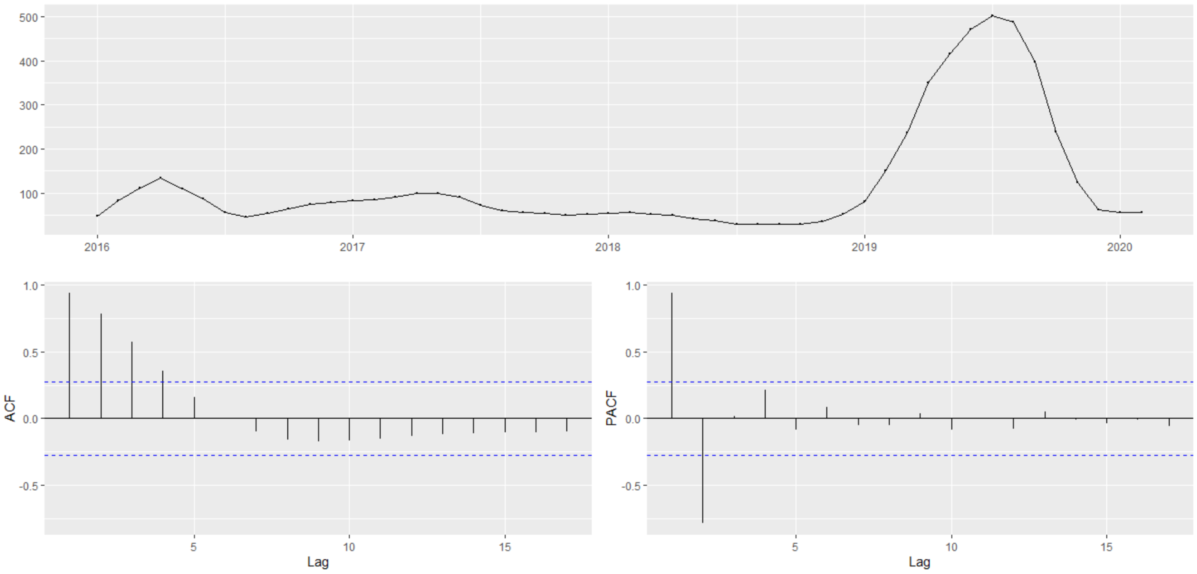

**Supplementary Figure 18. Residual plots of ARIMA of Hepatitis A incidences**

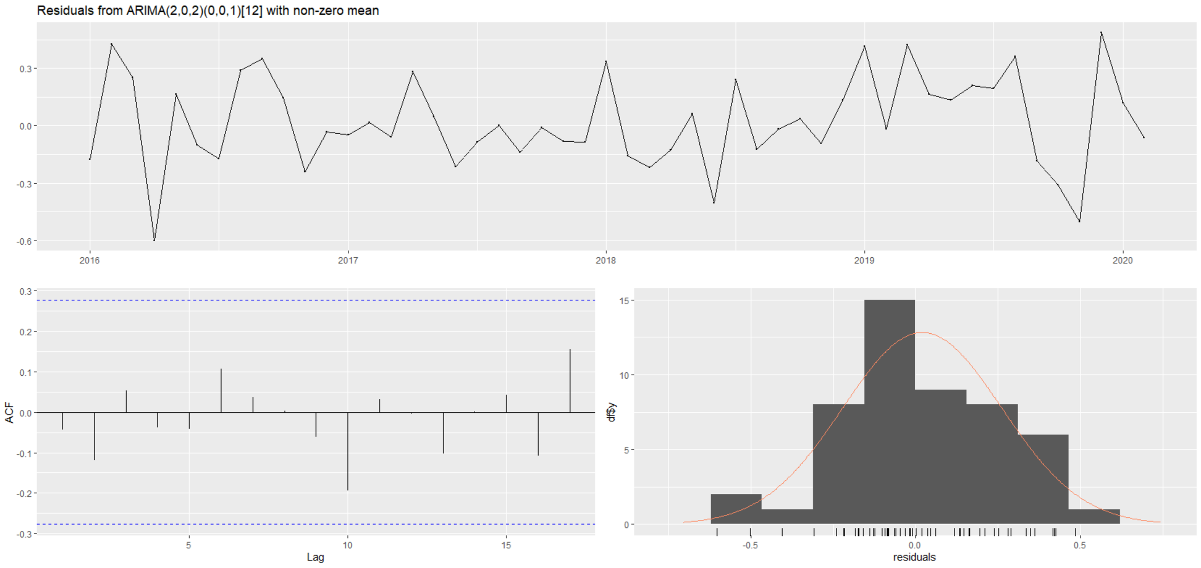

### Supplementary Figure 19. ACF and PACF plots of Enterohemorrhagic *E. coli* incidences

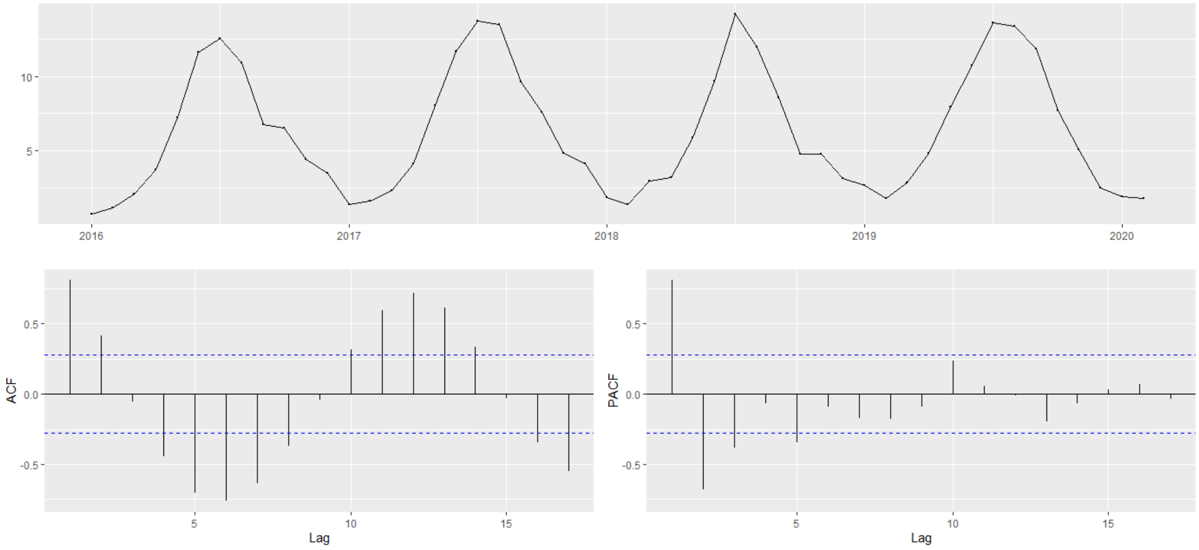

### Supplementary Figure 20. Residual plots of ARIMA of Enterohemorrhagic *E. coli* incidences

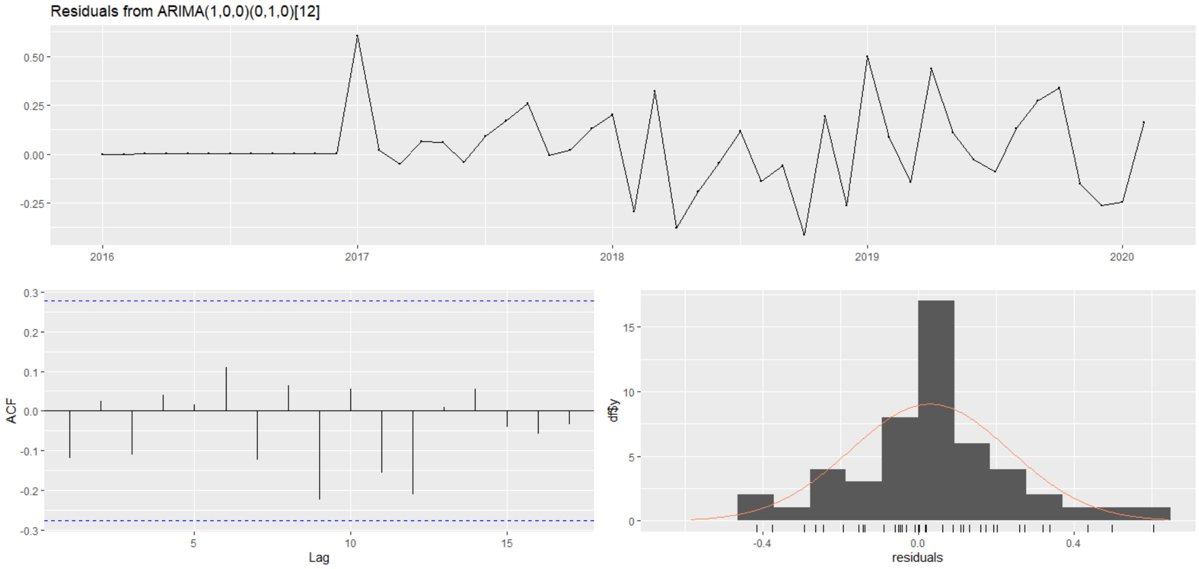

**Supplementary Figure 21. ACF and PACF plots of Bovine tuberculosis incidences**

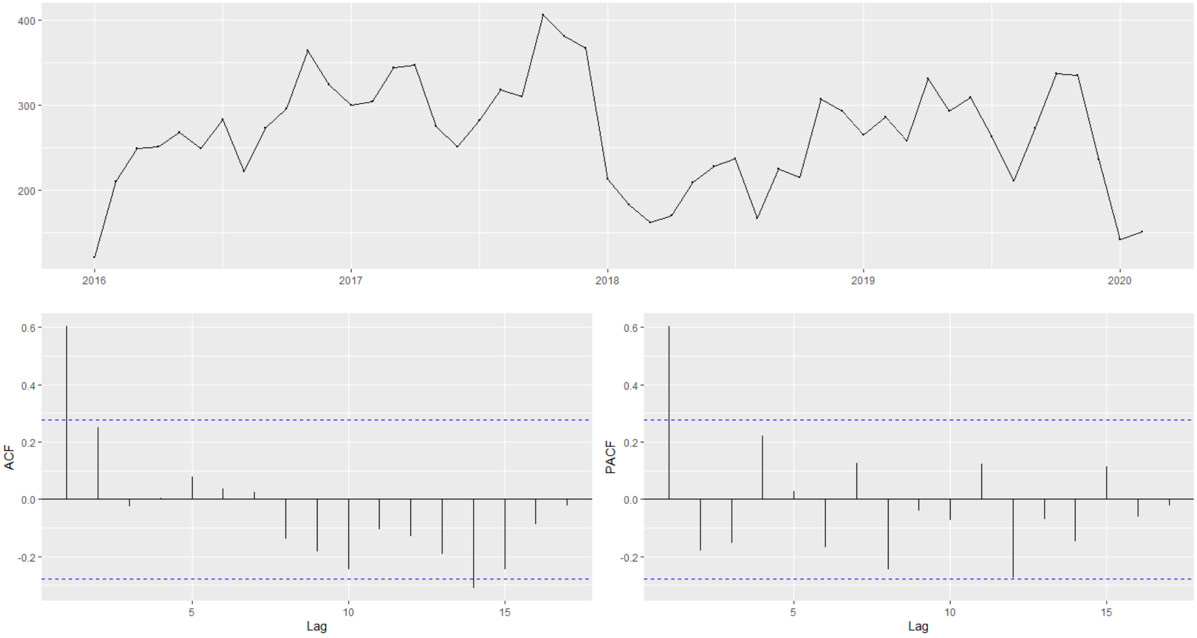

**Supplementary Figure 22. Residual plots of ARIMA of Bovine tuberculosis incidences**

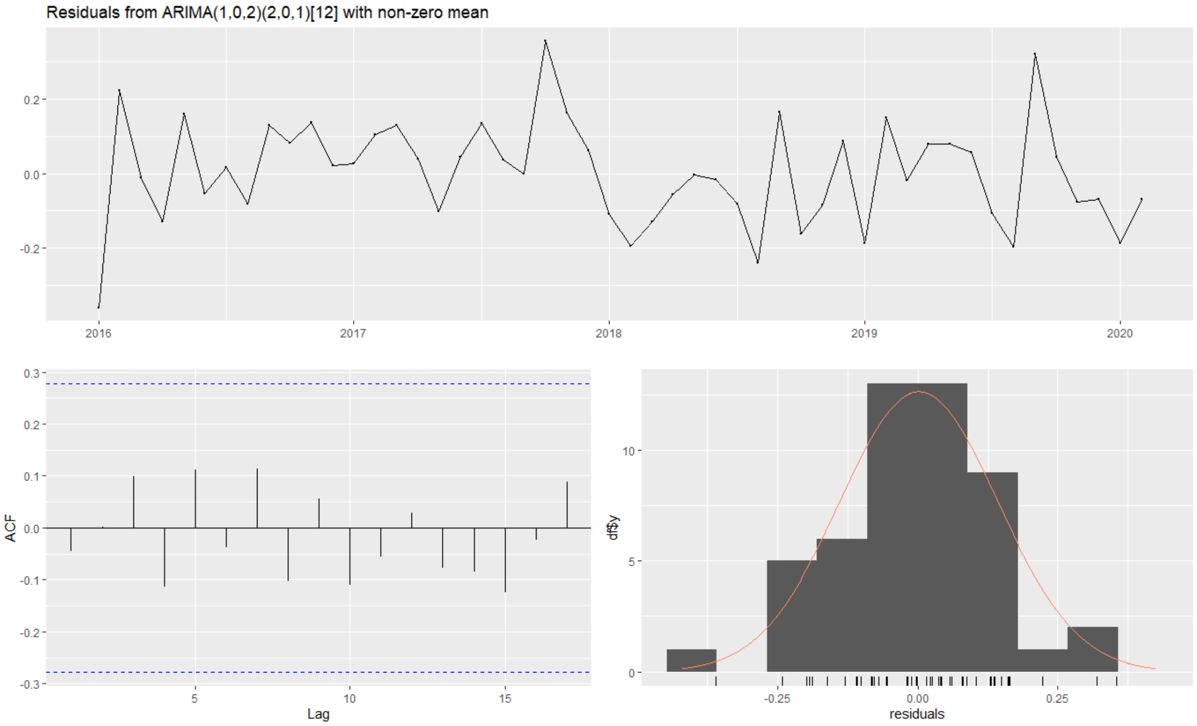

### Supplementary Figure 23. ACF and PACF plots of Bovine brucellosis incidences

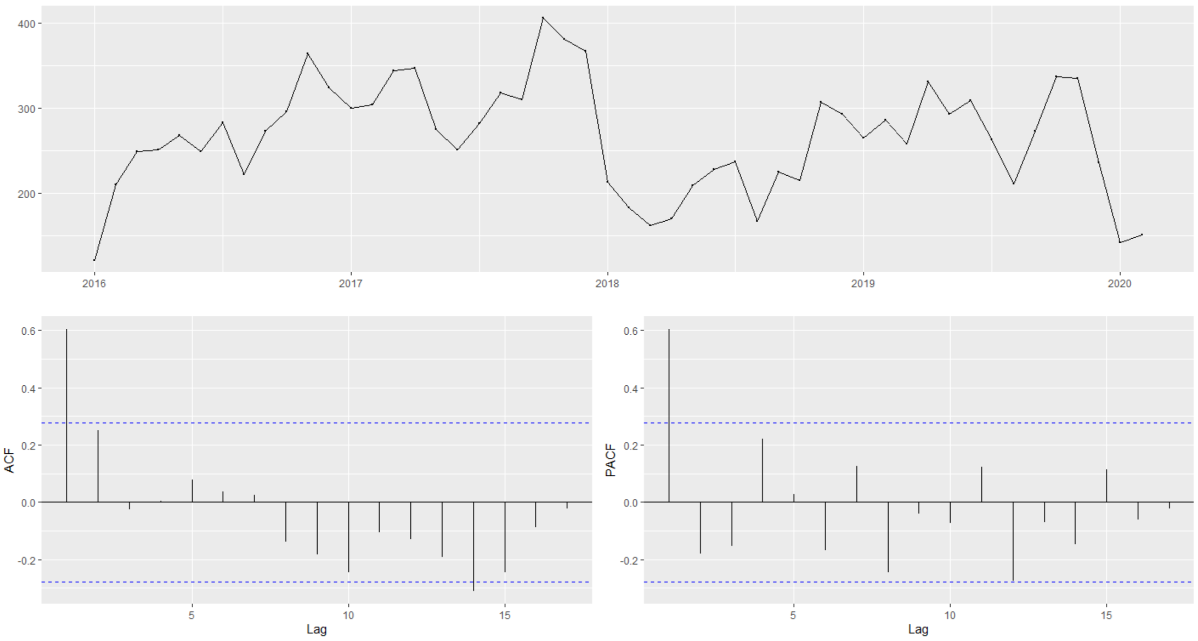

### Supplementary Figure 24. Residual plots of ARIMA of Bovine brucellosis incidences

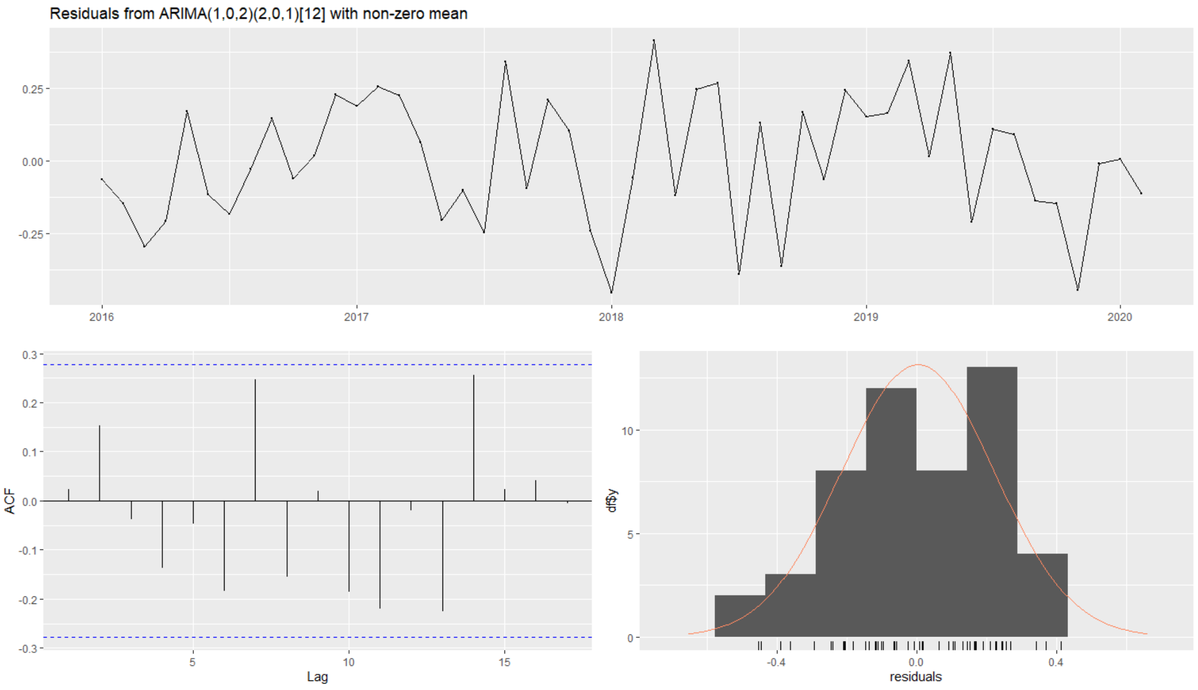
